# Sleep Temporal Entropy as a Digital Biomarker of Sleep Fragmentation for Cardiometabolic and Mortality Risk

**DOI:** 10.1101/2025.06.04.25328946

**Authors:** Jiong Chen, Clémence Cavaillès, Haoqi Sun, Haoran Zhao, Yaqing Gao, Donglin Xie, Xuesong Chen, Weijun Huang, Katie Stone, Hongliang Yi, Shenda Hong, Song Gao, Yue Leng

## Abstract

**Background:** Sleep fragmentation is increasingly recognized as a risk factor for cardiometabolic disease and mortality. However, existing measures primarily capture sleep–wake transitions and do not account for fragmentation within specific sleep stages, limiting their clinical utility.

**Methods:** We developed Sleep Temporal Entropy (STE), an entropy-based metric derived from hypnogram data to quantify overall and stage-specific sleep fragmentation. We evaluated its performance in two independent cohorts: a clinical cohort of 3,219 adults and a community-based cohort of 4,862 adults. Machine learning models and survival analyses were used to assess associations with cardiometabolic outcomes and mortality.

**Results:** STE outperformed conventional fragmentation metrics in predicting cardiometabolic conditions, including hypertension, diabetes, and hyperlipidemia. Across outcomes, STE-derived features consistently ranked among the top contributors in machine learning models. In longitudinal analyses, both low and high levels of STE were associated with increased mortality risk, forming a U-shaped relationship (p for nonlinearity = 0.025). This pattern was most pronounced for rapid eye movement (REM) sleep, where individuals in the lowest quintile of REM STE had increased risk of all-cause mortality (HR 1.58, 95% CI 1.16–2.15) and cardiovascular mortality (HR 2.83, 95% CI 1.66–4.80) compared with the reference group.

**Conclusions:** STE captures stage-specific sleep fragmentation and reveals its non-linear associations with health outcomes. These findings support the potential of STE as a scalable and interpretable biomarker for sleep health assessment and risk stratification.

**Plain Language Summary:** Sleep is important for health, but many people experience disrupted sleep without realizing its long-term effects. Scientists often measure sleep disruption by counting how often people wake up, but this does not fully describe how sleep organizes across the whole night and within different sleep stages. In this study, we developed a new way to measure interruptions in sleep, called Sleep Temporal Entropy. This method captures how sleep stages are distributed. We tested it in two large groups of people and used statistical and computer-based methods to study its relationship with health. We found that highly consolidated sleep and highly fragmented sleep patterns are linked to higher health risks, including risk of death and heart disease. People with a moderate level of sleep stability had the lowest risk. These findings suggest that healthy sleep is not just about fewer interruptions, but also about maintaining a stable and balanced sleep structure. This new measure may help improve how sleep is monitored and how future health risks are identified.

## Introduction

Sleep is essential for overall health and plays a key role in metabolic regulation, cognitive function, and immune defense^1–10^. Frequent sleep interruptions, known as sleep fragmentation, disrupt restorative sleep stages, compromising sleep quality and increasing risks for metabolic and cardiovascular conditions such as impaired glucose metabolism, hypertension, and cardiovascular disease (CVD) ^7,11–17^. Traditional measures of sleep fragmentation, such as Wake After Sleep Onset (WASO), sleep efficiency (SE), sleep fragmentation index (SFI), and arousal index (ArI), primarily quantify the frequency or duration of disruptive events. While informative for global sleep continuity, these metrics do not fully capture the micro-structural dynamics of transitions between sleep stages, limiting their ability to characterize the complexity of sleep architecture^18–25^.

Recent advances in digital sleep biomarkers such as arousal burden ^26^ and odds ratio product (ORP) ^27^ rely on electroencephalography (EEG) signals to characterize sleep continuity or fragmentation. Similarly, advanced metrics like Conditional Entropy (CE), Walsh Spectral Entropy (WSE), and Haar Spectral Entropy (HSE) capture unpredictability in sleep architecture within the frequency domain^24,28^. These approaches provide valuable insights into sleep-stage dynamics but often rely on high-resolution EEG signals and are not readily scalable for long-term or wearable-based monitoring^29^. Moreover, most studies have focused on correlations with existing sleep metrics or isolated associations with health outcomes, with limited direct comparison against conventional measures in predicting clinically relevant endpoints^21–24,28,30,31^. As a result, the incremental value of these metrics for risk stratification remains unclear.

To address these limitations, we introduce Sleep Temporal Entropy (STE), an entropy-based digital sleep biomarker derived from hypnogram data. STE quantifies fragmentation across both sleep–wake and stage-to-stage transitions by capturing the temporal distribution of sleep stage durations. Unlike conventional metrics that focus on event frequency, STE reflects both the occurrence of fragmentation and the stability of sleep stages, providing a more comprehensive representation of sleep architecture.

The objective of this study was to develop and evaluate STE as a novel metric of sleep fragmentation and to determine its association with cardiometabolic outcomes and mortality. We hypothesize that both excessively low and high STE values reflect dysregulated sleep-state transitions—characterized by either reduced flexibility or excessive instability—suggesting a non-linear (U-shaped) relationship with adverse health outcomes^32–34^. Conversely, entropy during wake periods (Wake STE), which primarily captures the frequency and stochasticity of awakenings, is expected to exhibit a monotonic association with increased risk.

In this study, we show that STE captures sleep fragmentation beyond conventional metrics and provides improved predictive value for cardiometabolic outcomes. STE consistently ranks among the most influential features in machine learning models and outperforms traditional fragmentation indices. Furthermore, STE demonstrates non-linear (U-shaped) associations with mortality, with both low and high entropy levels linked to increased risk. These findings establish STE as a scalable and interpretable digital biomarker that reflects the balance between sleep-stage stability and fragmentation, with important implications for risk stratification and sleep health assessment.

## Methods

### Study Design Overview

We used data from two independent cohorts: the Shanghai Sleep Health Study Cohort (SSHSC), a clinic-based population, and the Sleep Heart Health Study (SHHS), a community-based longitudinal cohort. In SSHSC, machine learning with post-hoc interpretability was applied to assess the contribution of STE relative to conventional sleep metrics in predicting cardiometabolic outcomes. In SHHS, we examined associations between STE and long-term outcomes, including all-cause and cardiovascular mortality, using both machine learning and survival analyses. Across both cohorts, we integrated statistical and interpretable machine learning approaches to evaluate whether STE captures information beyond traditional sleep fragmentation metrics, improves risk prediction, and reveals non-linear relationships with adverse health outcomes.

### Metrics Calculation

STE was derived from hypnogram data by segmenting each sleep stage into discrete episodes and quantifying the distribution of stage durations using Shannon entropy^32,35^:

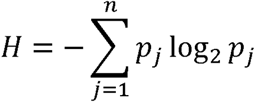

where *n* is the number of discrete episodes of a given sleep stage, and *p_j_* represents the proportion of total stage duration accounted for by segment *j* (***_j_*** = **duration***_j_*/∑**duration**).

Under this framework, entropy reflects the temporal dispersion of stage durations, with lower values indicating consolidated sleep and higher values indicating fragmented architecture.

STE was calculated for each sleep stage (N1, N2, N3, REM, and Wake) as well as for the entire sleep period (Overall STE). Detailed derivations and implementations of STE and other metrics are provided in the Supplementary Information (Mathematical and Statistical Methods). Code is publicly available at: https://github.com/JonChen916.

### Study Cohort Study Populations

The SSHSC is a clinic-based cohort of Chinese adults (≥18 years) presenting with snoring, who underwent overnight polysomnography (PSG) at the Sleep Center of Shanghai Sixth People’s Hospital between February 2017 and January 2022. Detailed recruitment procedures have been described previously^36–38^. Participants were excluded if they had severe systemic disease (e.g., cardiac, hepatic, pulmonary, or renal failure), severe psychiatric disorders or malignancy, or missing PSG data. Of 3,689 eligible individuals, 470 were excluded due to outlier values in total sleep time, sleep latency, or sleep efficiency, resulting in a final analytic sample of 3,219 participants. The study was approved by the institutional ethics committee (Approval No: 2019-KY-050[K]) and registered at the Chinese Clinical Trial Registry (ChiCTR1900025714; registration date: 6 September 2019). Written informed consent was obtained from all participants.

The SHHS is a large, community-based cohort investigating the relationship between sleep-disordered breathing and cardiovascular outcomes^39,40^. The study enrolled 6,441 adults aged ≥40 years between 1995 and 1998, with baseline assessments including questionnaires, anthropometry, and unattended overnight PSG. After exclusion of 637 participants from the Strong Heart Study and additional exclusions for missing cardiovascular mortality data (n = 760) and implausible STE values (<1; n = 182), 4,862 participants were included in the present analysis. SHHS data were obtained from the National Sleep Research Resource (NSRR; https://sleepdata.org/) in May 2024. All participating institutions obtained ethical approval, and all participants provided written informed consent.

### Exposures and Outcomes

Sleep fragmentation metrics were categorized into three groups: (1) traditional indices, including WASO, SE, and ArI; (2) established entropy-based measures, including CE, WSE and HSE; and (3) transition-based and temporal entropy metrics, including transition entropy (TE), semi-Markov entropy (SME), and STE. Detailed definitions and calculation procedures are provided in the Supplementary Information (Mathematical and Statistical Methods).

In the SSHSC cohort, cardiometabolic outcomes included hypertension, diabetes, and hyperlipidemia, defined based on clinical diagnosis and biochemical measurements. In the SHHS cohort, outcomes included all-cause mortality and CVD mortality. All-cause mortality was ascertained through follow-up interviews, medical records, and linkage to national death registries, while CVD mortality was determined based on adjudicated clinical data, including myocardial infarction, stroke, and coronary revascularization.

### Post-hoc Explanation in Machine Learning

In the SSHSC cohort, machine learning models were developed to evaluate the relative contribution of sleep fragmentation metrics to cardiometabolic outcomes, including hypertension, diabetes, and hyperlipidemia. XGBoost was used as the primary modeling approach due to its strong performance in structured data, with five-fold cross-validation applied to ensure model stability and generalizability. To assess the robustness of findings, additional models—including Random Forest, Support Vector Machine (SVM), K-Nearest Neighbors (KNN), and Logistic Regression—were implemented for comparison. Model performance was evaluated using area under the receiver operating characteristic curve (ROC AUC), F1-score, and precision–recall (PR) curves to account for class imbalance. Feature contributions were quantified using SHapley Additive exPlanations (SHAP), enabling both global ranking of feature importance and local interpretation of model predictions^41,42^. SHAP summary plots and dependence plots were used to characterize the directionality and potential non-linear effects of STE and other features. Model development was designed to enable stable and comparable estimation of feature importance, rather than to maximize predictive performance, thereby supporting interpretability of the incremental value of STE.

The same modeling framework was applied to the SHHS cohort for predicting all-cause and cardiovascular mortality. Models were constructed using both whole-night and stage-specific STE metrics to assess the incremental value of entropy-based measures. SHAP dependence plots were further used to examine interactions between STE and age.

### Statistical Analysis

To evaluate whether STE captures information distinct from conventional sleep metrics, Pearson correlation analyses were performed between STE and traditional indices, including WASO, ArI, and SE. Metrics were grouped into predefined classes (traditional, entropy-based, and novel measures) for structured comparison, and results were visualized using heatmaps. Given the large sample sizes in both the SSHSC and SHHS cohorts, Pearson correlation was used to assess linear relationships, and distributions of key variables were examined to confirm the robustness of the analysis.

In the SHHS cohort, Cox proportional hazards models were used to examine associations between sleep fragmentation metrics and all-cause and CVD mortality. Sleep metrics included SE, ArI, WASO, Overall STE, and stage-specific STE (Wake, N1–N3, REM, and NREM). Metrics were categorized into quintiles, with the third quintile (Q3) as the reference. Proportional hazards assumptions were tested for all models. Three models were constructed: Model 1 (unadjusted), Model 2 (adjusted for demographic variables including age, sex, race, and BMI), and Model 3 (fully adjusted for comorbidities, lifestyle factors, and sleep-related covariates, including total sleep time, smoking, sleep medication use, and apnea–hypopnea index). For stage-specific STE, the corresponding stage duration was included as an additional covariate. Sensitivity analyses were performed by further adjusting for hypoxic burden and T90. Multicollinearity was assessed using variance inflation factors (VIF), with all values below 5. Kaplan–Meier curves were generated to visualize survival differences across quintiles of entropy-based metrics. To assess potential non-linear associations, restricted cubic spline (RCS) models with four knots were fitted within the Cox regression framework, using the same covariates as the fully adjusted models. Hazard ratios (HRs) and 95% confidence intervals (CIs) were estimated, and non-linearity was evaluated using analysis of variance. To complement these analyses, SHAP dependence plots were used to visualize the relationship between STE and outcomes at the individual level and to explore interactions with age.

### Statistics and Reproducibility

All analyses were conducted using R (version 4.2.1) and Python (version 3.9). Statistical tests were two-sided, and exact p values are reported where applicable. Machine learning models were implemented using five-fold cross-validation to ensure stability and generalizability. All analyses were applied consistently across cohorts. Code used for analysis is publicly available.

## Results

### Visualizing Sleep Architecture Across Different Ranges of STE

To facilitate interpretation, we briefly outline the derivation of STE. STE is computed from hypnogram data by segmenting sleep-stage episodes and applying Shannon entropy to the normalized distribution of episode durations (*H* = − ∑*p_j_* log_2_ *p_i_*), where p_j_ denotes the proportion of each segment relative to the total time within a given stage. Unlike conventional linear metrics, STE captures both variability and temporal irregularity of stage durations. Higher STE reflects increased fragmentation and unpredictability of sleep-stage architecture, indicative of frequent transitions, whereas lower STE indicates more consolidated and stable stage organization with reduced stage cycling. We computed both whole-night STE and stage-specific STEs (e.g., REM, N3) for detailed analysis. A visual overview of the STE workflow is shown in Fig. 1a, with formal derivations provided in the Methods and Supplementary Information.

**Figure 1.**
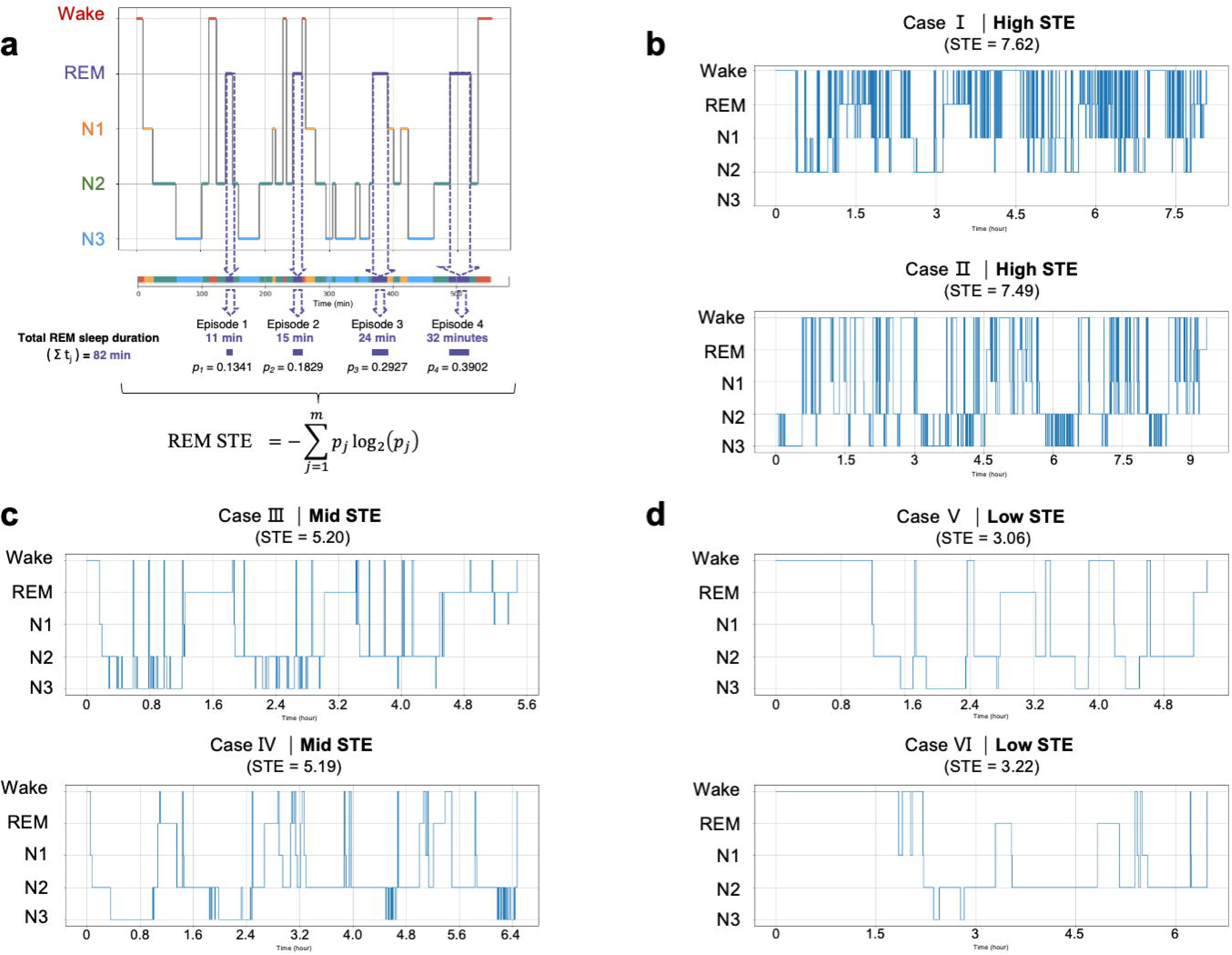
Concept of STE and illustrative examples across the STE spectrum. **a.** Computational framework of STE. Continuous durations of a specific sleep stage (illustrated here using REM sleep) are extracted from the hypnogram and segmented into discrete episodes. The proportion of each episode is calculated relative to the total duration of that stage, and Shannon entropy is applied to quantify the temporal distribution of stage durations. Higher STE values reflect greater dispersion of episode durations and increased fragmentation. **b–d.** Representative hypnograms across the STE spectrum. **b**, High STE, characterized by frequent stage transitions and short, fragmented sleep episodes; **c**, Intermediate STE, showing moderate variation in stage durations; **d**, Low STE, characterized by longer continuous episodes and fewer transitions. These examples illustrate differences in the temporal organization of sleep-stage dynamics across individuals.

To illustrate its interpretability, representative hypnograms spanning high, intermediate, and low STE values are shown in Fig. 1b–d. High STE cases (7.62 and 7.49) exhibited frequent stage transitions and marked fragmentation. Mid-range STE (5.19 and 5.20) showed moderate variability while preserving relatively complete sleep cycles. In contrast, low STE cases (3.22 and 3.06) appeared more consolidated but were often characterized by prolonged stage persistence or delayed sleep initiation, indicating that low STE does not necessarily reflect optimal sleep. These examples highlight the ability of STE to capture sleep architecture beyond conventional metrics.

### Participant characteristics

The demographic and clinical characteristics of the two study populations showed major differences (Table 1 and Table 2). Participants in the SSHSC (n=3,219) were younger (mean age: 41.0 years, SD=13.8) and more likely to be male (78.7%) compared to the SHHS cohort (n=4,862; mean age: 64.1 years, SD=11.3; male: 46.2%). Cardiometabolic disorders, including hypertension (32.7% vs. 27.6%) and diabetes (7.3% vs. 10.6%), were more common in the SHHS cohort compared to the SSHSC cohort.

**Table 1:**
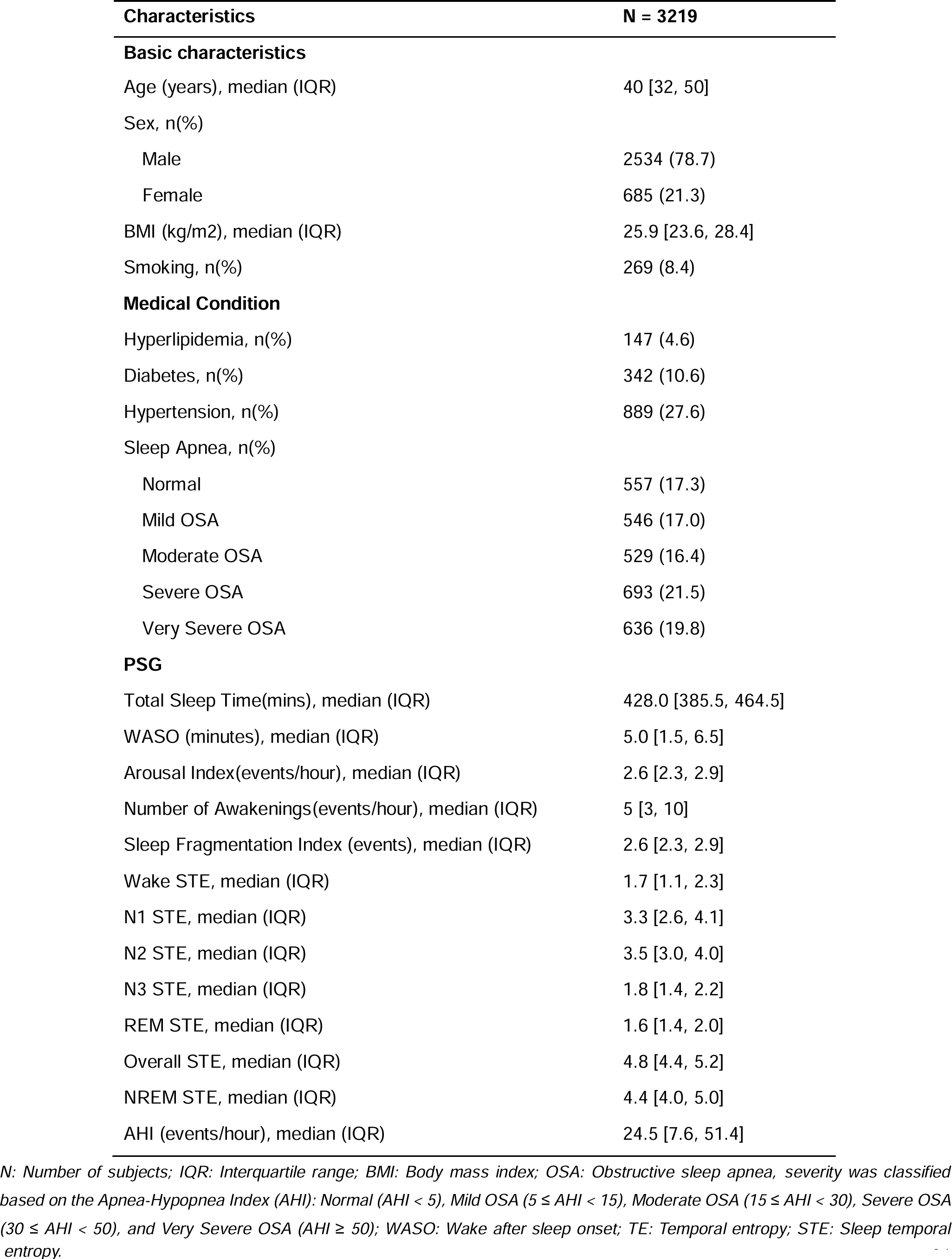
Population characteristics of the Shanghai Sleep Health Study Cohort.

**Table 2:**
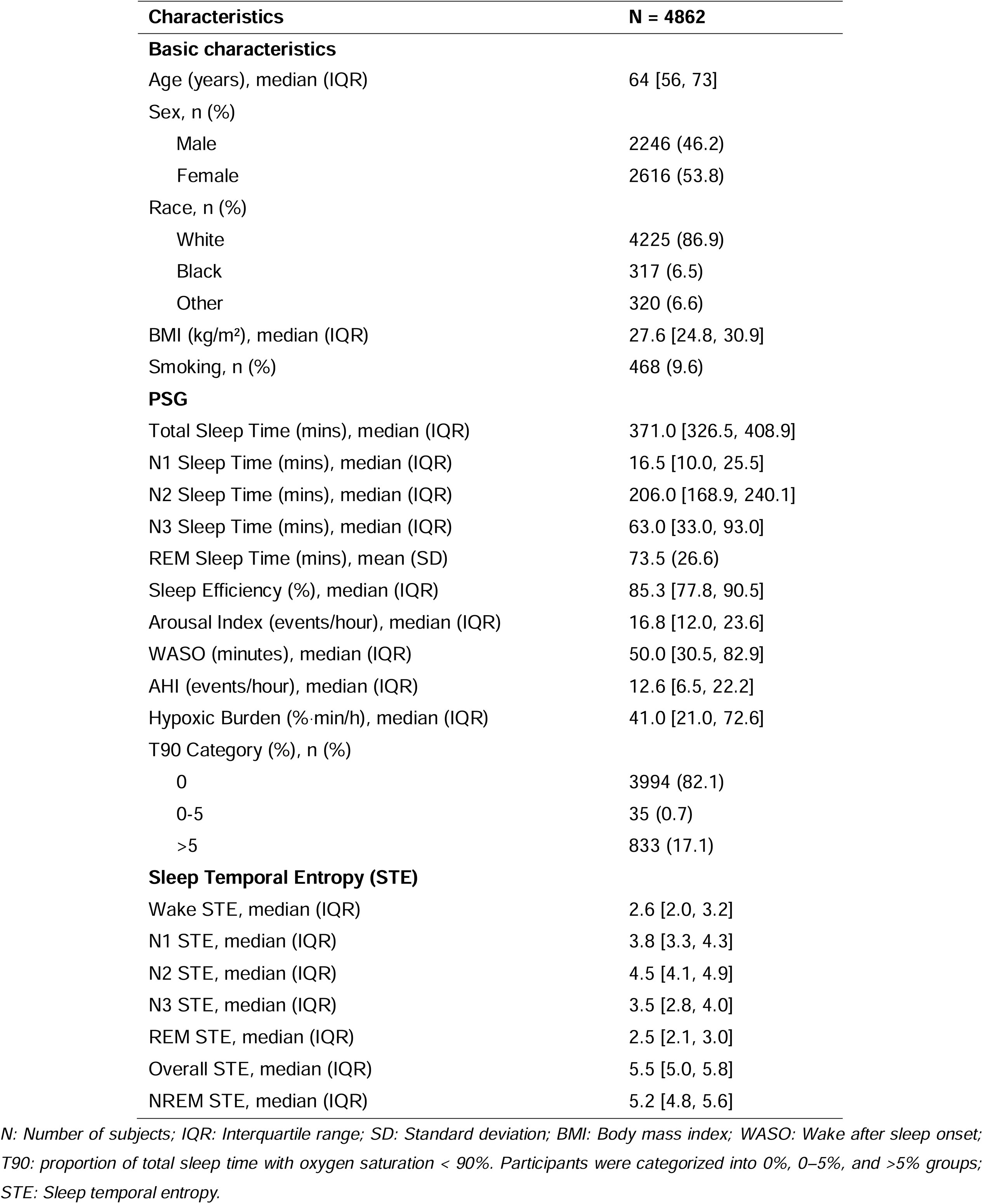
Cohort characteristics of the Sleep Heart Health Study Characteristics N = 4862.

### STE captures distinct dimensions of sleep fragmentation and predicts cardiometabolic outcomes

We first examined whether STE captures aspects of sleep architecture that are not represented by conventional fragmentation metrics. Correlation analyses revealed strong inter-correlations among traditional measures (e.g., WASO, sleep efficiency, ArI), reflecting their shared sensitivity to global sleep disruption. In contrast, entropy-based markers—including STE and related measures—formed a distinct cluster with moderate internal correlations but weak associations with traditional indices. For instance, WASO showed minimal correlation with stage-specific STE (e.g., N3 STE: r = –0.06; REM STE: r = 0.03), indicating that conventional metrics primarily quantify macro-level sleep disruption, whereas STE captures stage-specific temporal dynamics of sleep architecture. To validate these findings in an independent cohort, we next evaluated STE in the SHHS (Fig. 3a). Consistent with the SSHSC cohort, both whole-night and stage-specific STE measures showed similar patterns in relation to conventional sleep fragmentation metrics. Supporting analyses are provided in Supplementary Figure 3 and Supplementary Table 2, with additional results from the SSHSC cohort shown in Supplementary Figure 1.

**Figure 3.**
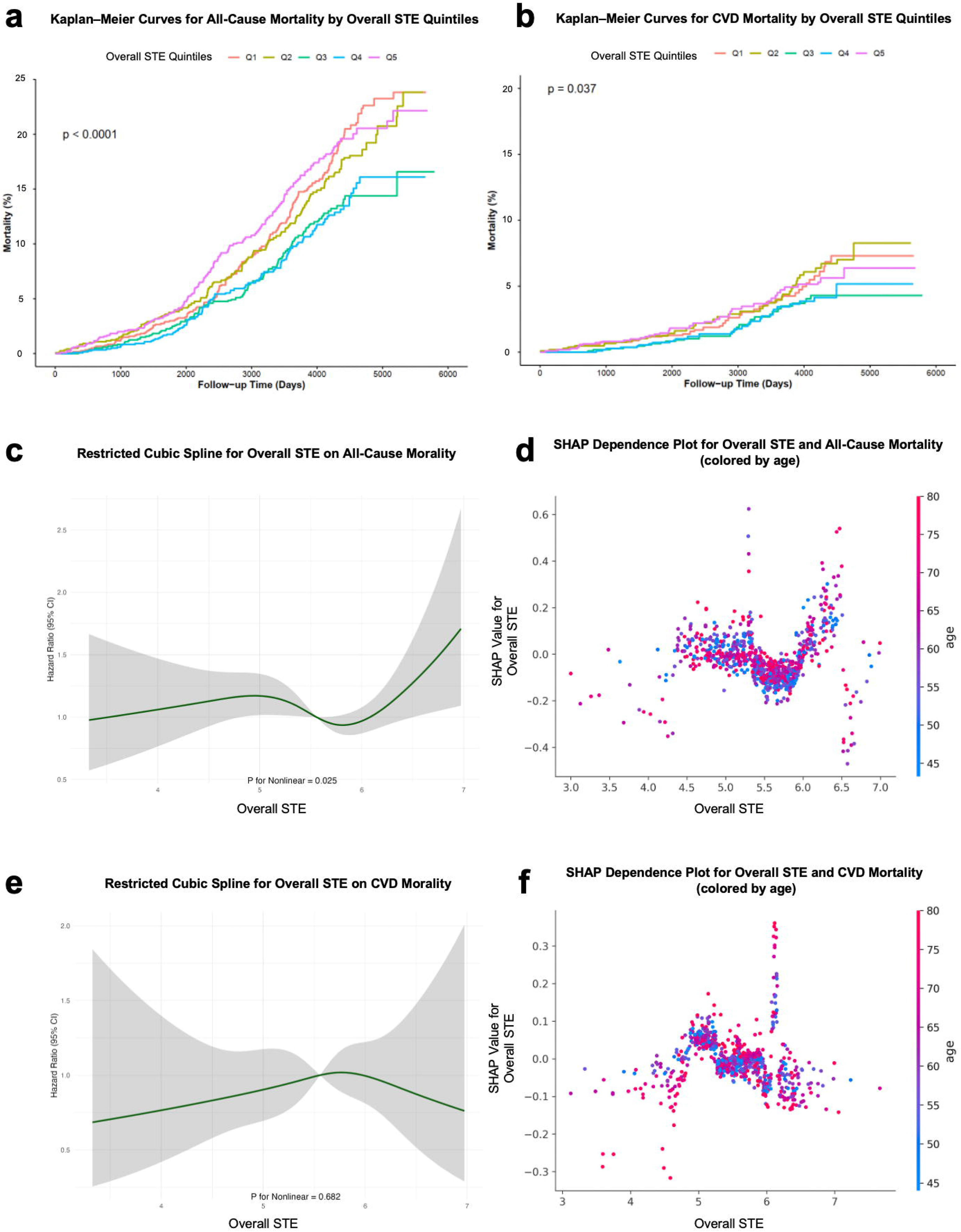
Evaluation of sleep fragmentation metrics for mortality prediction in the SHHS. **a.** Correlation heatmap of sleep fragmentation metrics. **b.** ROC curves for five machine learning models predicting CVD mortality. **c-d**. SHAP feature importance rankings for all-cause mortality (c) and CVD mortality (d) using overall sleep metrics. **e-f**. SHAP feature importance rankings for all-cause (e) and CVD (f) mortality after incorporating stage-specific STE metrics, highlighting their additional predictive contribution.

To further illustrate this distinction, we compared hypnograms of two representative individuals with nearly identical macro-sleep profiles (WASO, ArI, NoA, SFI) (Fig. 2a). Despite comparable conventional metrics, their underlying sleep-stage transition patterns differed markedly. STE, particularly stage-specific STE, effectively distinguished these differences and revealed heterogeneous instability across sleep stages, highlighting its ability to resolve temporal dynamics of sleep architecture that remain latent in traditional indices.

**Figure 2.**
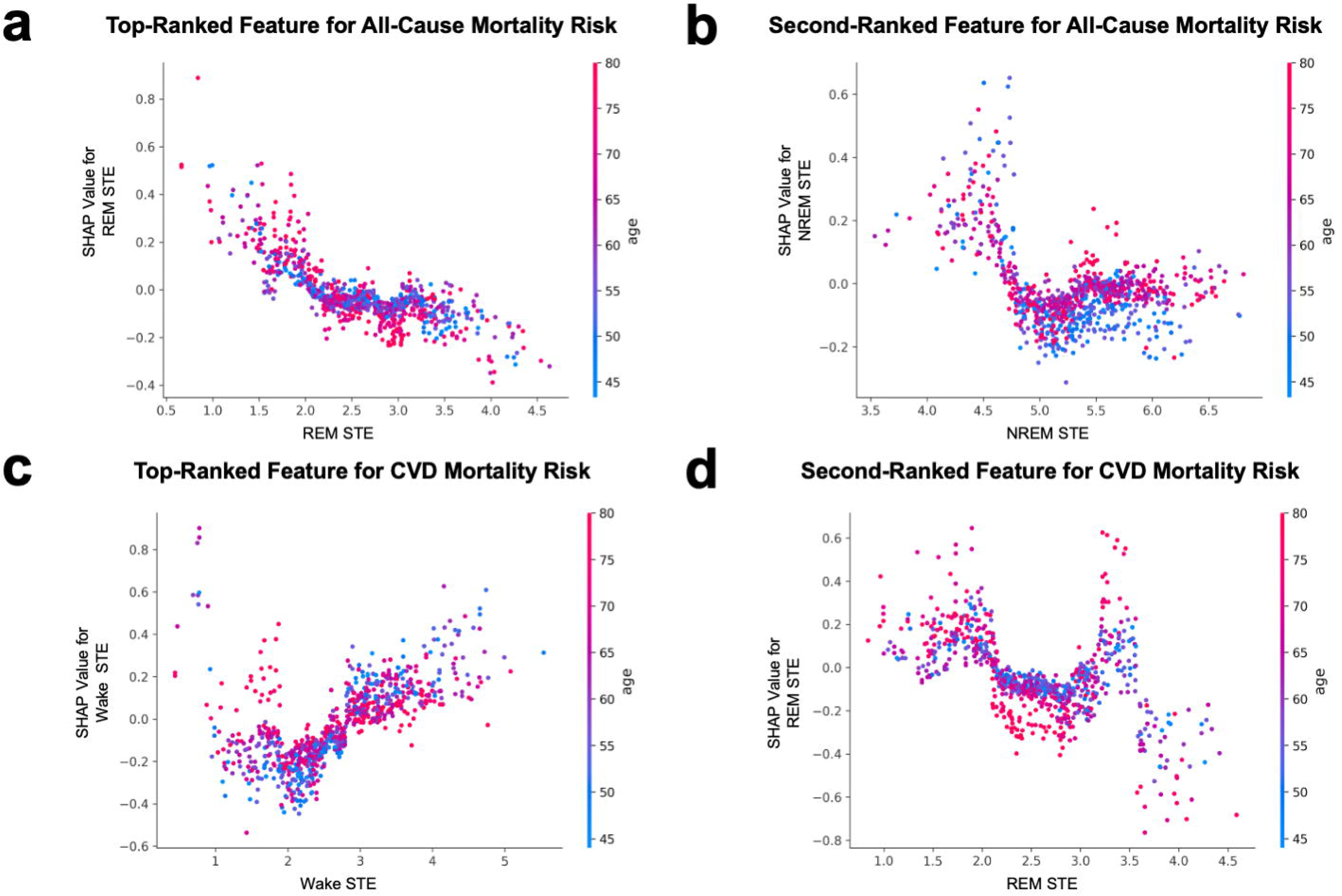
Predictive performance of sleep fragmentation metrics for cardiometabolic disorders. **a.** Case comparison of two participants with similar macro-sleep metrics (WASO and ArI) but divergent STE profiles. Hypnograms and radar charts illustrate temporal differences not captured by conventional metrics. **b.** SHAP summary plots for hypertension, diabetes, and hyperlipidemia. Features are ranked by mean absolute SHAP values; each dot represents an individual observation, with color indicating feature value (red, high; blue, low). **c.** Integrated feature importance rankings for whole-night (left) and stage-specific (right) models across all outcomes. STE-derived metrics (hatched bars) consistently rank among the top predictors, often outperforming conventional fragmentation indices and clinical variables (e.g., BMI, age).

We next assessed the contribution of STE to cardiometabolic outcomes using machine learning models. Model performance was evaluated using ROC AUC and F1-score, with PR analyses performed to account for class imbalance (Supplementary Figure 2 and Supplementary Table 1). Across all outcomes (hypertension, diabetes, hyperlipidemia), STE-derived metrics consistently ranked among the top contributing predictors in SHAP analyses (Fig. 2b). Within sleep-related features, stage-specific STE measures (e.g., REM STE, N3 STE) showed particularly strong contributions, outperforming conventional fragmentation metrics. Notably, in low-prevalence outcomes such as hyperlipidemia, the STE-based framework achieved a PR-AUC of 0.62 with a recall of 0.998, indicating high sensitivity to fragmentation-related signals. Across all models, STE demonstrated greater contribution than both conventional fragmentation metrics and previously proposed entropy-based measures (Fig. 2b-c), underscoring its robustness as a predictive feature.

### STE demonstrates robust and consistent importance across machine learning frameworks

To ensure that feature importance rankings were not model-dependent, we evaluated multiple machine learning algorithms. For both all-cause and CVD mortality, XGBoost and Random Forest achieved the highest performance, with XGBoost reaching an accuracy of 0.870 and ROC AUC of 0.836 for all-cause mortality, and 0.952 and 0.838 for CVD mortality. SVM also performed competitively, while KNN consistently underperformed (Supplementary Figure 4, Table 3 and 4). Two sets of models were compared: one using whole-night (non–stage-specific) features and another incorporating stage-specific STE. Their predictive performance was comparable (Supplementary Table 5). However, when comparing models with identical covariates, inclusion of STE modestly improved performance metrics, supporting its incremental predictive value.

**Table 3:**
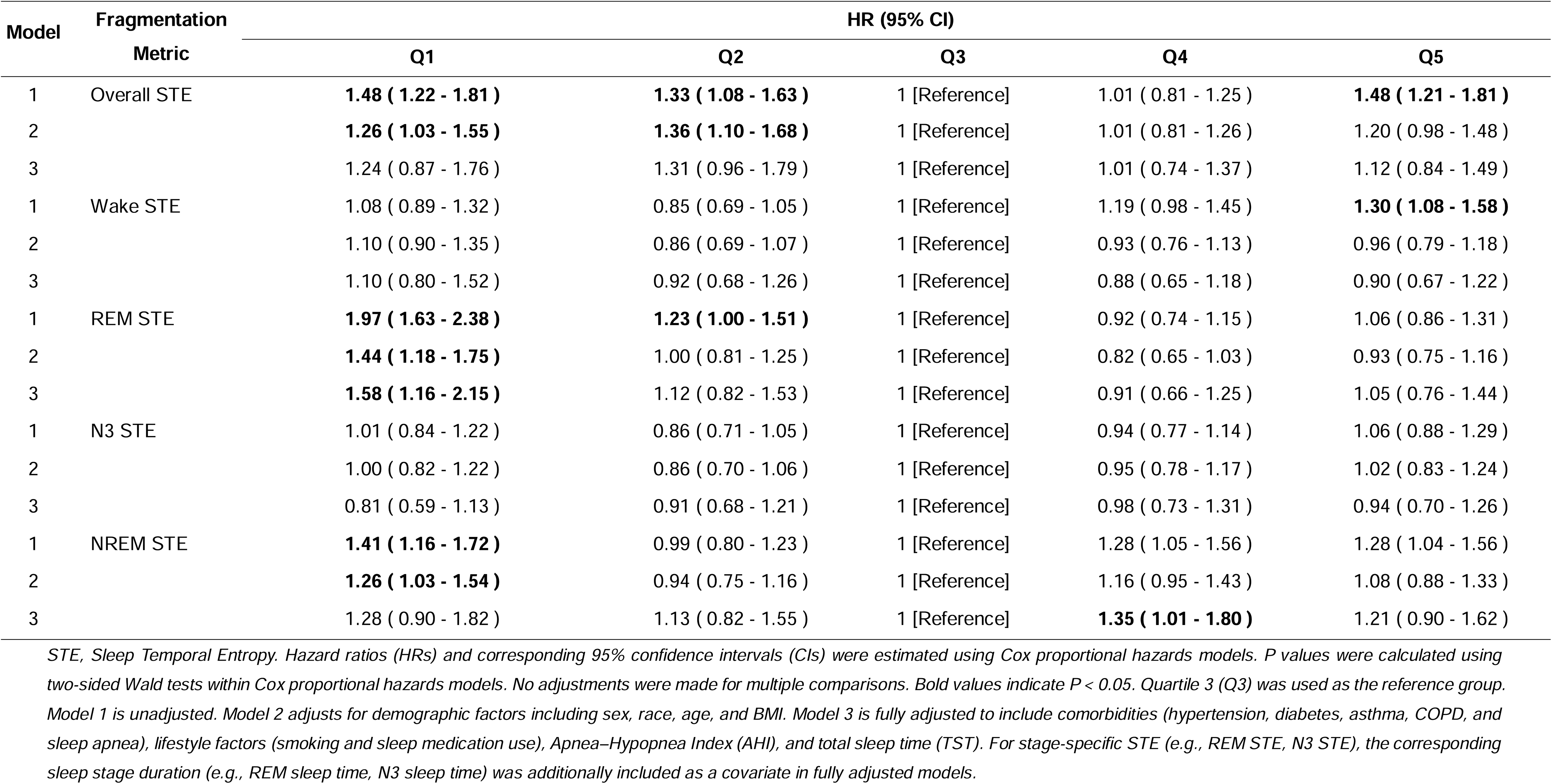
Hazard Ratios for All-Cause Mortality Across Sleep Temporal Entropy Model Fragmentation.

SHAP analyses further demonstrated that STE retained high feature importance across independent datasets (Fig. 3c–f). For all-cause mortality, age remained the dominant predictor, followed by Overall STE and total sleep time. In stage-specific models, REM STE and NREM STE emerged as key contributors. Similar patterns were observed for CVD mortality. Despite class imbalance in CVD mortality (∼6%), PR curve analyses showed that STE-based models substantially outperformed baseline models (Supplementary Figure 5 and 6), reinforcing the robustness of STE-derived signals.

### STE is associated with long-term mortality risk with non-linear patterns

Participants who died were older and had a higher prevalence of cardiometabolic conditions compared with those who survived (Supplementary Table 6 and 7). In longitudinal analyses, STE was significantly associated with both all-cause and cardiovascular mortality. Proportional hazards assumptions were tested for all Cox models, with results provided in Supplementary Table 8 and Figure 7, supporting the validity of the models. In Cox regression models (Tables 3 and 4), REM STE showed the strongest and most consistent associations. Compared to the reference group (Q3), individuals in the lowest quintile (Q1) of REM STE had increased risk of all-cause mortality (HR = 1.97, 95% CI: 1.63–2.38), which remained significant after full adjustment (HR = 1.58, 95% CI: 1.16–2.15). Elevated risk was also observed in the highest quintile. Similar patterns were found for CVD mortality. Kaplan–Meier curves demonstrated clear separation across STE quintiles (Supplementary Figure 8, 9 and 10), with intermediate ranges (Q3–Q4) showing the lowest mortality risk (Fig. 4a–b). These associations remained robust after additional adjustment for hypoxic burden and T90 (Supplementary Table 9, 10 and 11).

**Figure 4.**
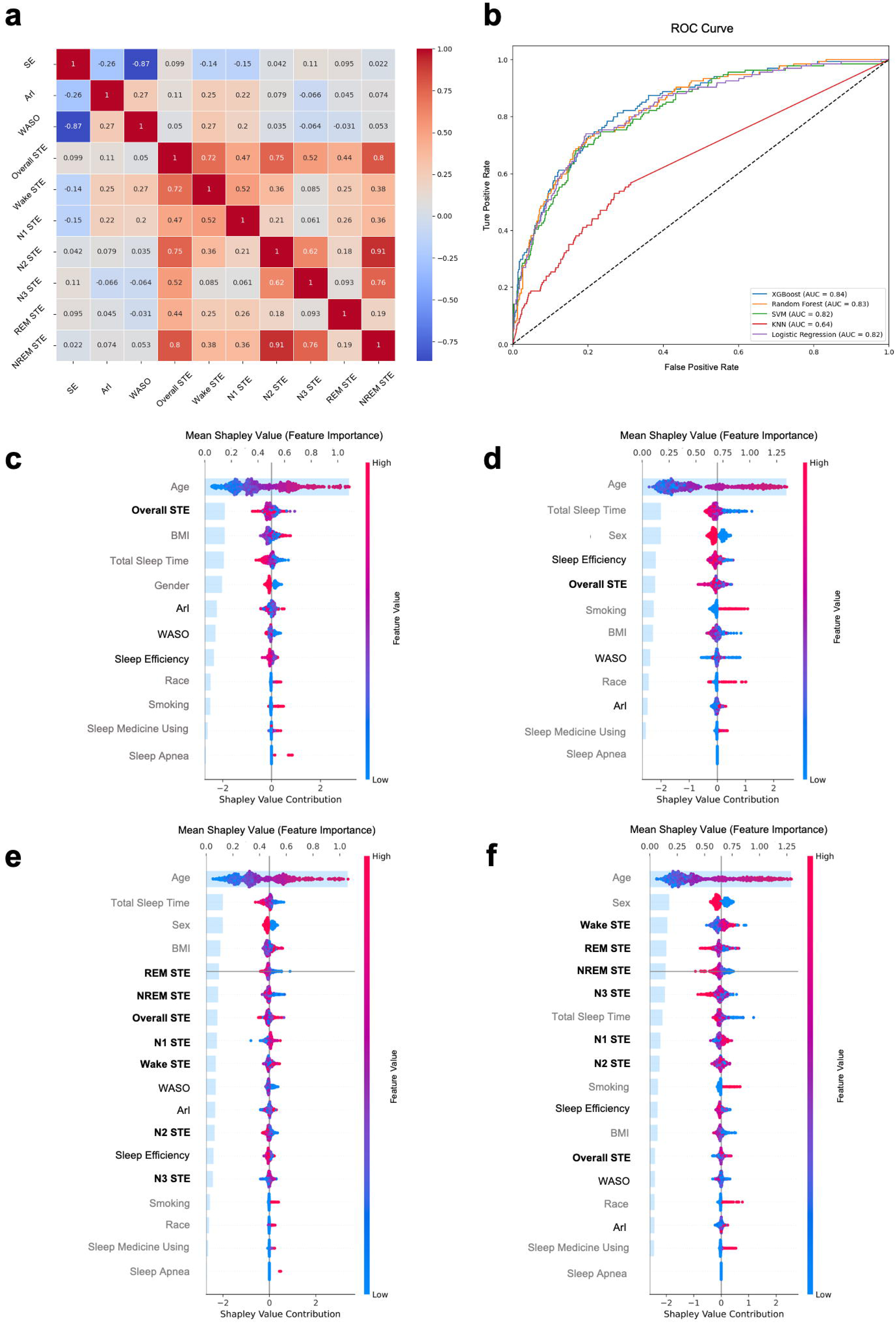
Association of overall STE with all-cause and CVD mortality in the SHHS. **a-b**. Kaplan–Meier curves for all-cause (a) and CVD (b) mortality across quintiles of overall STE. P values were calculated using two-sided log-rank tests (all-cause mortality: P = 0.000096; CVD mortality: P = 0.036941). No adjustments were made for multiple comparisons. **c-e**. RCS analyses of overall STE with all-cause (c) and CVD (e) mortality. Hazard ratios were estimated using Cox proportional hazards models with restricted cubic splines, and shaded areas represent 95% confidence intervals (CIs). P values for nonlinearity were calculated using Wald tests (two-sided), with no adjustment for multiple comparisons. **d-f**. SHAP age dependence plots for overall STE in relation to all-cause (d) and CVD (f) mortality, illustrating age-modulated effects.

**Table 4:**
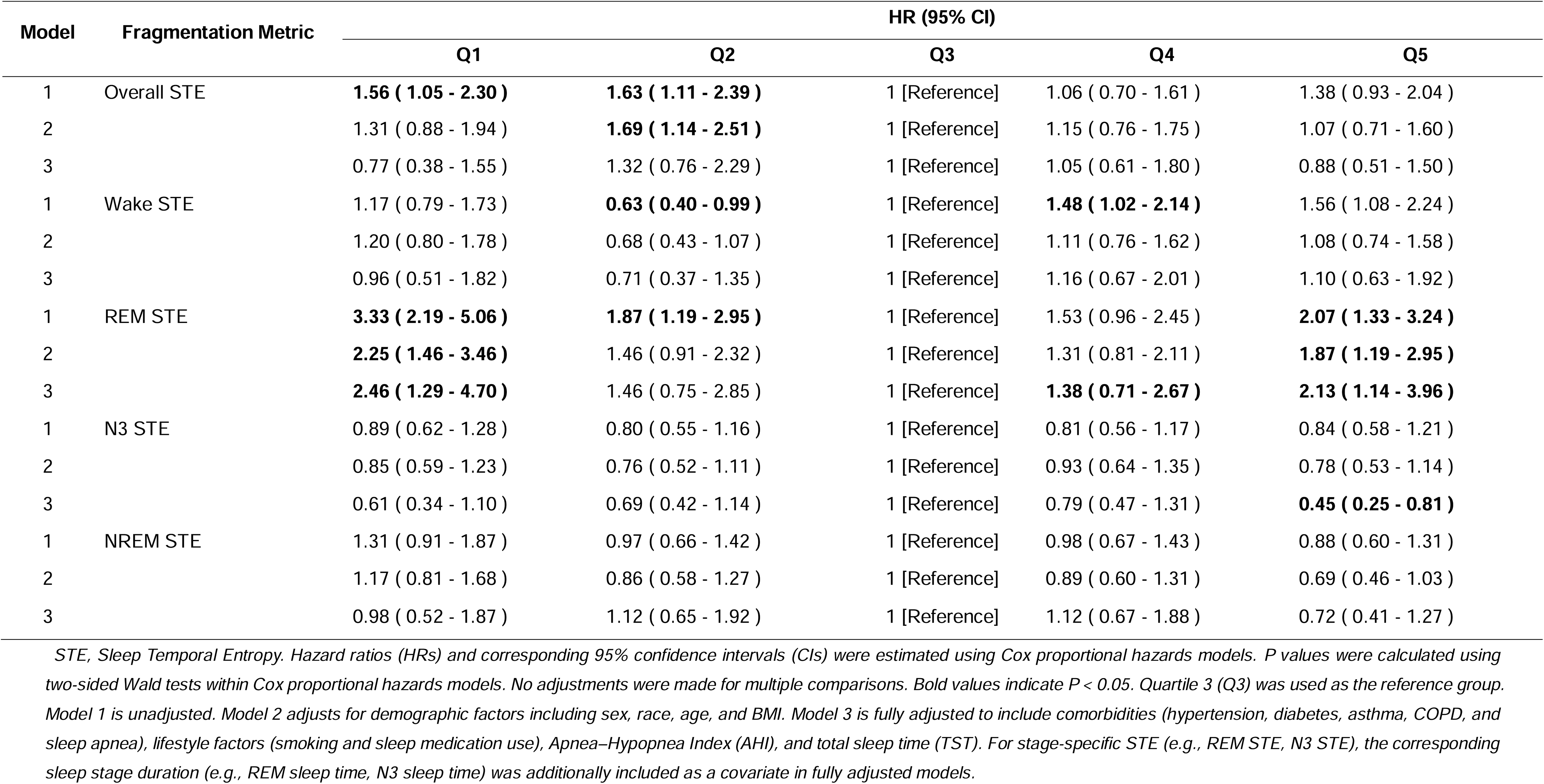
Hazard Ratios for CVD Mortality Across Sleep Temporal Entropy.

To further characterize these relationships, we applied RCS models and SHAP dependence analyses (Supplementary Figure 11 and 12). RCS results showed a significant U-shaped association between Overall STE and all-cause mortality (p for nonlinearity = 0.025), whereas no significant non-linearity was observed for CVD mortality (Fig. 4c, e). SHAP dependence plots corroborated these findings, revealing a consistent U-shaped pattern across the distribution of STE values (Fig. 4d, f). Age modified this relationship, with older individuals exhibiting higher risk at comparable STE levels.

Analysis of stage-specific STE further demonstrated similar non-linear patterns. Dependence plots showed that both low and high values of REM, NREM, and N3 STE were associated with increased SHAP values, indicating elevated predicted risk (Fig. 5). These patterns were also observed across cardiometabolic outcomes (Supplementary Figure 13-18), although effect sizes varied. Notably, REM STE exhibited the most consistent and pronounced non-linear associations across both cross-sectional and longitudinal analyses, supporting its important role in linking sleep architecture to health outcomes.

**Figure 5.**
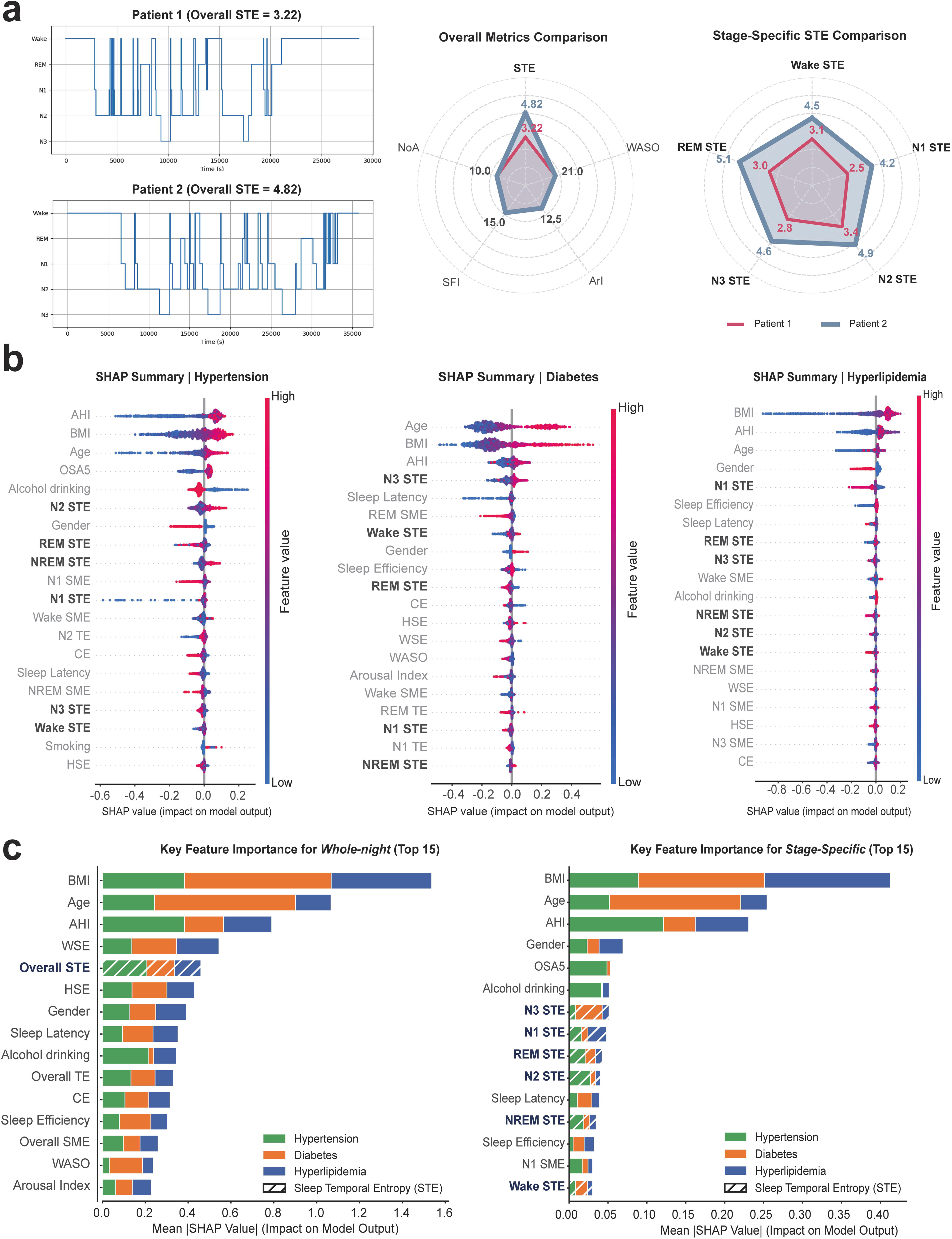
Age dependence of top-ranked STE features for mortality risk. **a-b**. SHAP age dependence plots for the top STE features in all-cause mortality; **c**, **d**, corresponding plots for CVD mortality.

## Discussion

In this study, we developed and validated a entropy-based digital sleep biomarker to quantify sleep fragmentation and characterize abnormalities in sleep-stage temporal dynamics across both clinical and community-based populations. By integrating machine learning with statistical analyses, we demonstrated that STE captures both whole-night fragmentation and stage-specific instability, and is associated with cardiometabolic outcomes and mortality across diverse settings. Consistent with our hypothesis, we observed a U-shaped relationship between STE and adverse health outcomes, with both low and high entropy values associated with increased risk, while intermediate levels appeared protective. This pattern was evident for overall and several stage-specific STE measures, but not for Wake STE. Specifically, in the SSHSC cohort, U-shaped associations were observed for N3 STE with hypertension, REM STE with diabetes, and N2/N3 STE with hyperlipidemia. In the SHHS cohort, mortality outcomes were associated with Overall STE, NREM STE, and REM STE, with REM STE showing the most consistent and robust associations across models.

This U-shaped pattern may reflect two complementary mechanisms. First, from a sleep-cycle perspective, healthy sleep typically consists of 4–6 NREM–REM cycles per night^43,44^. Reduced STE may reflect insufficient transitions and incomplete cycling, indicative of impaired sleep dynamics. Second, among individuals with comparable cycle counts, increased fragmentation leads to higher STE, reflecting instability in sleep architecture, which has been linked to adverse physiological outcomes^45–48^. Together, these mechanisms suggest that STE captures a balance between cycle integrity and fragmentation, with an optimal range corresponding to stable and physiologically effective sleep organization^49,50^.

Our findings extend prior work on entropy-based sleep metrics by demonstrating a non-linear association with health outcomes. Previous entropy measures, such as WSE or HSE, primarily quantify signal-level unpredictability and often show linear relationships with sleep disruption^24,28^. In contrast, STE captures the temporal distribution of stage durations at the hypnogram level, enabling sensitivity to both insufficient transitions and excessive fragmentation. Compared with transition-based entropy measures (e.g., transition entropy and semi-Markov entropy), which focus primarily on state-to-state transition probabilities, STE explicitly captures the temporal distribution, duration, and organization of sleep stage episodes over the course of the night, thereby providing a more comprehensive representation of sleep-stage dynamics. This distinction may explain why STE uniquely exhibits U-shaped associations, identifying an “optimal range” of sleep-stage dynamics that is not captured by conventional or spectral entropy measures. Our results are consistent with recent findings by Bechny et al. and align with studies showing that stage-transition dynamics carry prognostic information for cardiovascular risk^51,52^. Importantly, our findings also reconcile prior literature reporting predominantly linear associations between sleep fragmentation and adverse outcomes^15,20,24,26,53,54^. Traditional metrics primarily capture sleep–wake transitions, which correspond to Wake STE in our framework. We similarly observed a monotonic relationship between Wake STE and risk, supporting the interpretation that wake-related fragmentation follows a linear pattern, whereas stage-specific fragmentation exhibits non-linear dynamics.

Not all outcomes demonstrated clear U-shaped patterns, particularly for metabolic conditions. This is consistent with prior literature showing that associations between sleep characteristics and metabolic outcomes are less consistent than those observed for mortality^55,56^. In addition, the analyses of metabolic outcomes in this study were cross-sectional, whereas mortality analyses were longitudinal. Cross-sectional associations reflect concurrent relationships and are more susceptible to confounding, whereas longitudinal analyses capture long-term risk and temporal relationships. Finally, metabolic outcomes are strongly influenced by dominant risk factors such as BMI and lifestyle behaviors, which may attenuate or obscure the contribution of sleep-related measures^53,57,58^. Together, these factors may explain why non-linear patterns were less apparent for metabolic outcomes in this study.

Stage-specific analyses further highlight the physiological relevance of different sleep stages. REM STE consistently ranked among the top predictors across cardiometabolic and mortality outcomes and showed the most robust associations in survival analyses. These findings align with prior evidence linking REM sleep disruption to mortality and metabolic dysfunction^59–62^. Notably, our results extend this literature by demonstrating that both insufficient transitions into REM (low REM STE) and excessive fragmentation within REM (high REM STE) are associated with increased risk, suggesting that both continuity and dynamic regulation of REM sleep are critical for health. Similarly, N3 (slow-wave sleep) plays a well-established role in metabolic regulation^63–65^. Our findings suggest that fragmentation within N3, as captured by N3 STE, may also contribute to adverse outcomes, although the U-shaped pattern was less pronounced in older populations such as SHHS. This attenuation may reflect age-related changes in sleep architecture and the dominant influence of other risk factors.

Our study has several strengths. We quantified sleep fragmentation at the level of stage transitions rather than relying solely on sleep–wake metrics, integrated machine learning with survival analysis for consistent validation, directly compared STE with established fragmentation metrics, and demonstrated reproducibility across heterogeneous populations. By combining SHAP-based interpretability with traditional statistical methods, our study provides a complementary framework for understanding complex, non-linear relationships between sleep architecture and health outcomes. Compared to RCS analyses, which are sensitive to sample size at distribution extremes, SHAP dependence plots offer a more granular, individual-level perspective on risk patterns, supporting their use in future biomarker evaluation^66^.

Several limitations should be considered. First, STE was derived from manually scored hypnograms rather than raw EEG signals and therefore depends on the accuracy of sleep stage classification. Sleep staging is subject to inter-scorer variability and may be influenced by signal quality, scoring criteria, and algorithmic differences, which could introduce measurement error into entropy estimates. Although STE captures the overall temporal organization of sleep and may be relatively robust to minor misclassifications, such variability may still limit its precision. However, manual PSG-based scoring remains the clinical gold standard, and most established sleep fragmentation metrics are likewise derived from hypnogram annotations. Aligning STE with this reference framework enhances comparability with prior work and supports clinical interpretability. Future studies may benefit from incorporating automated, AI-based sleep staging approaches to improve scalability and consistency across large datasets. In addition, while existing wearable devices currently have limited accuracy for sleep staging, STE does not require raw EEG signals and may remain informative even under simplified staging, making it potentially compatible with future improvements in wearable sleep technologies. Second, although STE demonstrated robust feature importance, overall predictive performance was insufficient for direct clinical application. This likely reflects the multifactorial nature of cardiometabolic diseases, in which sleep represents only one of several contributing domains. Third, the contribution of sleep fragmentation to cardiometabolic outcomes is modest relative to established risk factors such as BMI, which may attenuate the apparent importance of STE in predictive models. Accordingly, STE should be interpreted as a complementary marker of sleep fragmentation rather than a primary risk factor. Nevertheless, STE consistently demonstrated greater importance than conventional sleep metrics, suggesting that it captures additional aspects of sleep architecture. Fourth, the calculation of overall STE assumes equal contributions across sleep stages, which may oversimplify underlying physiological dynamics. Finally, we did not exclude participants using sleep-related or other medications, but instead adjusted for medication use in the analyses; residual confounding may therefore remain. This approach preserves cohort representativeness and may enhance generalizability to real-world populations, although generalizability may still be limited by the absence of younger and healthier populations.

In summary, STE is a scalable and interpretable digital biomarker that captures the temporal dynamics of sleep-stage transitions beyond conventional fragmentation metrics. By revealing non-linear associations between sleep fragmentation and health outcomes, STE provides a new perspective on sleep architecture and its role in cardiometabolic and mortality risk. These findings support the potential utility of STE for risk stratification and for advancing the understanding of sleep-related health mechanisms.

## Data availability

The data used in this study include the Shanghai Sleep Health Study Cohort and the Sleep Heart Health Study.

The Shanghai Sleep Health Study Cohort data are not publicly available due to privacy and ethical restrictions involving human participants. Access to these data may be requested from the corresponding author, Yue Leng, subject to institutional review board approval and data use agreements. Requests should include a brief research proposal and will be reviewed within 2–4 weeks. Restrictions may apply to the use of the data in accordance with institutional policies governing human subject research.

The Sleep Heart Health Study data are publicly available from the National Sleep Research Resource repository (https://sleepdata.org/).

The source data underlying Figures 2a, 2b, 3b–f, and 4a–e are provided in Supplementary Data 1–7 (see individual file descriptions for detailed mappings). All other relevant data supporting the findings of this study are available from the corresponding author upon reasonable request.

## Code availability

The code used to compute sleep temporal entropy (STE) and perform the analyses in this study is publicly available at https://github.com/JonChen916.

All scripts necessary to reproduce the main results are included in the repository.

## Author Contributions

J.C. conceptualized the study, developed the methodology, performed the analysis, and drafted the manuscript.

C.C. contributed to methodological review and manuscript revision.

H.S. contributed to study conceptualization.

H.Z. contributed to both study conceptualization and methodological development.

Y.G. contributed to critical revision of the manuscript.

D.X. and X.C. curated and managed the data.

W.H. and H.Y. collected the data.

K.S. contributed to conceptual development and manuscript revision.

S.H. and S.G. contributed to study design and manuscript revision.

Y.L. supervised the study and oversaw manuscript revision.

All authors reviewed and approved the final version of the manuscript.

## Ethics Statement

All procedures involving human participants were conducted in accordance with the Declaration of Helsinki.

The Shanghai Sleep Health Study Cohort was approved by the Ethics Committee of Shanghai Sixth People’s Hospital Affiliated to Shanghai Jiao Tong University School of Medicine (Approval No: 2019-KY-050[K]). Written informed consent was obtained from all participants.

The Sleep Heart Health Study was approved by the institutional review boards of all participating institutions, including Brigham and Women’s Hospital, Boston University Medical Center, Case Western Reserve University, Johns Hopkins University, the University of Arizona, the University of California, Davis, the University of Minnesota, and the University of Washington. All participants provided written informed consent. Permission for secondary use of the data was obtained through the National Sleep Research Resource.

The current study involved secondary analysis of de-identified data. Ethical approval for this study was reviewed and approved by the Ethics Committee of Peking University. The requirement for additional informed consent was waived because all data were anonymized.

## Abbreviations

STE: Sleep Temporal Entropy
REM: rapid eye movement
NREM: non-rapid eye movement
WASO: wake after sleep onset
ArI: arousal index
SHAP: SHapley Additive exPlanations
ROC: receiver operating characteristic
CVD: cardiovascular disease
RCS: Restricted cubic spline.

## Acknowledgements

This study was additionally supported by the Ministry of Science and Technology of the People’s Republic of China (Grant No. 2021ZD0201902). The funding sources had no role in the design and conduct of the study; collection, management, analysis, and interpretation of the data; preparation, review, or approval of the manuscript; or the decision to submit the manuscript for publication.

The Sleep Heart Health Study (SHHS) was supported by National Heart, Lung, and Blood Institute cooperative agreements U01HL53916 (University of California, Davis), U01HL53931 (New York University), U01HL53934 (University of Minnesota), U01HL53937 and U01HL64360 (Johns Hopkins University), U01HL53938 (University of Arizona), U01HL53940 (University of Washington), U01HL53941 (Boston University), and U01HL63463 (Case Western Reserve University). The National Sleep Research Resource was supported by the National Heart, Lung, and Blood Institute (R24 HL114473, 75N92019R002).

The Shanghai Sleep Health Study Cohort (SSHSC) was supported by grants from the Ministry of Science and Technology of the People’s Republic of China (Grant Nos. 2021ZD0201900, 2021ZD0201902), Shanghai Municipal Commission of Science and Technology (Grant No. 18DZ2260200), Shanghai Science and Technology Innovation Program (Grant No. 20Y11902100), National Natural Science Foundation of China (Grant Nos. 82071030, 81700896, 81770988, 81970869), and Shanghai Shen-Kang Hospital Management Center (Grant Nos. SHDC2020CR2044B, SHDC2020CR3056B).

