## Supplementary Information for "Sleep Temporal Entropy as a Digital Biomarker of Sleep Fragmentation for Cardiometabolic and Mortality Risk"

### Mathematical and Statistical Methods

#### 1. Traditional Metrics

1.1 **Sleep Latency**

The time from lights out to the first onset of sleep (non-wake state), typically measured in minutes.

$$\text{Sleep Latency}=T_{\text{first sleep}}-T_{\text{lights out}}$$

Where $T_{\text{first sleep}}$  is the time of the first sleep onset, and $T_{\text{lights out}}$ is the time when lights are turned off.

1.2 **REM Latency**

The time from sleep onset to the first occurrence of REM (Rapid Eye Movement) sleep.

$$\text{REM Latency}=T_{\text{first REM}}-\text{Sleep Latency}$$

where $T_{\text{first REM}}$ is the time of the first REM stage.

1.3 **WASO，Wake After Sleep Onset**

The total duration of wakefulness occurring after sleep onset, typically measured in minutes.

$$\text{WASO}=\sum_{i=1}^{n} D_{\text{wake},i}$$

where $D_{\text{wake},i}$ represents the duration of each wake period, and the sum gives the total WASO.

1.4 **Arousal Index**

The number of awakenings per hour of sleep, used to measure the degree of sleep disruption.

$$\text{Arousal Index}=\frac{\text{Number of Awakenings}}{\text{Total Sleep Time (hours)}}$$

1.5 **Number of Awakenings**

The total number of awakenings that occur from sleep onset until final awakening. This metric is determined by counting the specific occurrences of awakenings during sleep.

1.6 **SFI，Sleep Fragmentation Index**

$$\text{SFI}=\frac{\text{Number of Awakenings}+\text{Stage Transitions}}{\text{Total Sleep Time (hours)}}$$

where $\text{Stage Transitions}$ refers to the number of transitions between different sleep stages.

#### 2. Entropy Based Metrics

2.1 **Walsh Spectral Entropy (WSE):**

A measure of the complexity of sleep stage transitions based on the Walsh transform. It reflects the unpredictability in the sleep stage transitions encoded into frequency components.

$$\text{WSE}=-\sum_{i=1}^{N} \left( \frac{\left| F_{\text{walsh}}\left( i \right) \right|^{2}}{\sum_{i=1}^{N} \left| F_{\text{walsh}}\left( i \right) \right|^{2}} \right)\log\left( \frac{\left| F_{\text{walsh}}\left( i \right) \right|^{2}}{\sum_{i=1}^{N} \left| F_{\text{walsh}}\left( i \right) \right|^{2}} \right)$$

where $F_{\text{walsh}}\left( i \right)$ represents the Walsh-transformed frequency components of the encoded sleep stages, and $N$ is the number of components.

2.2 **Haar Spectral Entropy (HSE):**

A measure of the complexity of sleep stage transitions based on the Haar wavelet transform. It reflects the unpredictability in the sleep stage transitions when decomposed into wavelet components.

$$\text{HSE}=-\sum_{i=1}^{N} \left( \frac{\left| C_{\text{haar}}\left( i \right) \right|^{2}}{\sum_{i=1}^{N} \left| C_{\text{haar}}\left( i \right) \right|^{2}} \right)\log\left( \frac{\left| C_{\text{haar}}\left( i \right) \right|^{2}}{\sum_{i=1}^{N} \left| C_{\text{haar}}\left( i \right) \right|^{2}} \right)$$

where $C_{\text{haar}}\left( i \right)$ represents the Haar wavelet coefficients of the encoded sleep stages, and $N$ is the number of components.

2.3 **Conditional Entropy (CE):**

A measure of the dependency between consecutive sleep stages, assessing the predictability of a future stage given the current stage. It quantifies the uncertainty in predicting the next sleep stage.

$$\text{CE}\left( L \right)=-\sum_{j=1}^{M} p_{j}\left( L \right)\log\left( p_{j}\left( L \right) \right)+\sum_{k=1}^{M-1} p_{k}\left( L-1 \right)\log\left( p_{k}\left( L-1 \right) \right)$$

Where $p_{j}\left( L \right)$ is the probability of occurrence of a pattern of $L$ consecutive sleep stages, and $p_{k}\left( L-1 \right)$ is the probability of occurrence of a pattern of $\left( L-1 \right)$ consecutive sleep stages.

#### 3. New Entropy Based Metrics

3.1 **Semi-Markov Transition Matrix Calculation:**

The Semi-Markov transition matrix extends the traditional Markov chain by incorporating the duration of time spent in each state before transitioning to another state.

**Step 1: Define States and Transitions**

- Let $S=\{s_{1},s_{2},\ldots,s_{n}\}$ be the set of sleep states (e.g., Wake, N1, N2, N3, REM).

- For each state $s_{i}$, we record the time spent in that state (stay time) and the transitions to other states $s_{j}$.

**Step 2: Transition Count Matrix**

- Construct a Transition Count Matrix $C$, where each element $C_{\mathrm{ij}}$ represents the number of observed transitions from state $s_{i}$ to state $s_{j}$.

$$C=\left( \begin{matrix} C_{11} & C_{12} & \cdots& C_{1n} \\ C_{21} & C_{22} & \cdots& C_{2n} \\ \vdots& \vdots& \ddots& \vdots\\ C_{n1} & C_{n2} & \cdots& C_{nn} \end{matrix} \right)$$

**Step 3: Calculate Stay Times**

- For each state $s_{i}$, we collect the Stay Times $T_{i}=\{t_{i1},t_{i2},\ldots,t_{\mathrm{im}}\}$ which are the durations spent in that state before transitioning to another state.

**Step 4: Fitted Distributions and Cumulative Distribution Function (CDF)**

- Use Kernel Density Estimation (KDE) to fit a distribution $f_{i}\left( t \right)$ to the stay times $T_{i}$.

- Calculate the Cumulative Distribution Function (CDF) $F_{i}\left( t \right)$ for the fitted distribution.

**Step 5: Semi-Markov Transition Probability**

- The semi-Markov transition probability from state $s_{i}$ to state $s_{j}$. is calculated by adjusting the standard Markov transition probability with the CDF of the stay time distribution:

$P_{ij}^{\text{semi-Markov}}=P_{ij}\times F_{i}\left( t \right)$

Where:

- $P_{ij}$ is the probability of transitioning from state $s_{i}$ to state $s_{j}$ as derived from the transition count matrix.

- $F_{i}\left( t \right)$ is the CDF value at the median stay time for state $s_{i}$.

**Step 6: Construct the Semi-Markov Transition Matrix**

- The Semi-Markov Transition Matrix $M$ is given by:

$$M=\left( \begin{matrix} P_{11}^{\text{semi-Markov}} & P_{12}^{\text{semi-Markov}} & \cdots& P_{1n}^{\text{semi-Markov}} \\ P_{21}^{\text{semi-Markov}} & P_{22}^{\text{semi-Markov}} & \cdots& P_{2n}^{\text{semi-Markov}} \\ \vdots& \vdots& \ddots& \vdots\\ P_{n1}^{\text{semi-Markov}} & P_{n2}^{\text{semi-Markov}} & \cdots& P_{nn}^{\text{semi-Markov}} \end{matrix} \right)$$

3.2 **Entropy Calculation Based on the Semi-Markov Matrix**

With the semi-Markov transition matrix $M$ computed, we calculate entropy to measure the unpredictability or randomness in the transitions.

**Overall Shannon Entropy for the Semi-Markov Matrix**

- The Overall Entropy $H\left( M \right)$ for the semi-Markov matrix is calculated as:

$$H\left( M \right)=-\sum_{i=1}^{n} \sum_{j=1}^{n} M_{ij}\log_{2} \left( M_{ij} \right)$$

Where:

- $M_{ij}$ is the element of the semi-Markov matrix representing the probability of transitioning from state $s_{i}$ to state $s_{j}$.

**State-Specific Entropy**

- The Entropy for Specific State $s_{i}$ is calculated by considering only the transitions originating from that state:

$$H\left( s_{i} \right)=-\sum_{j=1}^{n} M_{ij}\log_{2} \left( M_{ij} \right)$$

**NREM Entropy**

- NREM Entropy aggregates the entropy values of all NREM sleep stages (N1, N2, N3):

$$H_{\text{NREM}}=\sum_{s_{i}\in\{\text{N1},\text{N2},\text{N3}\}} H\left( s_{i} \right)$$

3.3 **Markov Transition Matrix Entropy**

For comparison, entropy can also be calculated using the standard Markov transition matrix $T$ (ignoring the time spent in each state):

- Overall Markov Entropy:

$$H\left( T \right)=-\sum_{i=1}^{n} \sum_{j=1}^{n} T_{ij}\log_{2} \left( T_{ij} \right)$$

- State-Specific Markov Entropy:

$$H_{T}\left( s_{i} \right)=-\sum_{j=1}^{n} T_{ij}\log_{2} \left( T_{ij} \right)$$

- NREM Markov Entropy:

$$H_{\text{NREM}}^{T}=\sum_{s_{i}\in\{\text{N1},\text{N2},\text{N3}\}} H_{T}\left( s_{i} \right)$$

3.4 **Overall Entropy of the Transition Matrix (T_Overall_Entropy)**

The "T_Overall_Entropy"*measures the overall unpredictability or randomness in the transitions between all sleep stages as represented by the Markov transition matrix $T$.

$$\text{T}\text{\_}\text{Overall}\text{\_}\text{Entropy}=-\sum_{i=1}^{n} \sum_{j=1}^{n} T_{ij}\log_{2} \left( T_{ij} \right)$$

Where:

- $T_{ij}$ is the probability of transitioning from state $s_{i}$ to state $s_{j}$ in the standard Markov transition matrix.

- The summation is taken over all possible transitions between the $n$ states.

3.5 **NREM Entropy of the Transition Matrix (T_NREM Entropy)**

The "T_NREM Entropy" measures the unpredictability or randomness of transitions specifically within the NREM sleep stages (N1, N2, N3) based on the Markov transition matrix $T$.

$$\text{T}\text{\_}\text{NREM Entropy}=\sum_{s_{i}\in\{\text{N1},\text{N2},\text{N3}\}} H_{T}\left( s_{i} \right)$$

Where:

- $(H_{T}\left( s_{i} \right)=-\sum_{j=1}^{n} T_{\mathrm{ij}}\log_{2} \left( T_{\mathrm{ij}} \right))$is the entropy for a specific state $s_{i}$ in the transition matrix $T$.

- The summation for $\text{T}\text{\_}\text{NREM Entropy}$ is taken over all NREM stages $s_{i}\in\{\text{N1},\text{N2},\text{N3}\}$

3.6 **State-Specific Temporal Entropy**

The Temporal Entropy for each sleep stage (e.g., Wake, N1, N2, N3, REM) measures the unpredictability or randomness in the distribution of time durations spent in that particular stage.

For a given sleep stage $s_{i}$, the Temporal Entropy is calculated as:

$$H_{\text{Temporal}}\left( s_{i} \right)=-\sum_{j=1}^{m} p_{j}\log_{2} \left( p_{j} \right)$$

Where:

- $p_{j}$ is the probability associated with the $j$-th duration spent in stage $s_{i}$.

- $p_{j}=\frac{t_{j}}{\sum_{k=1}^{m} t_{k}}$ , where $t_{j}$ is the $j$-th duration and $m$ is the total number of duration entries for stage $s_{i}$.

- The summation is over all temporal durations $t_{j}$ spent in stage $s_{i}$.

3.7 **Overall Temporal Entropy**

The Overall Temporal Entropy is a measure of the randomness in the distribution of time durations across all sleep stages combined.

$$H_{\text{Overall}\text{\_}\text{Temporal}}=-\sum_{i=1}^{n} p_{i}\log_{2} \left( p_{i} \right)$$

Where:

- $p_{i}$ is the probability associated with the $i$-th duration across all sleep stages.

- $p_{i}=\frac{t_{i}}{\sum_{j=1}^{n} t_{j}}$ , where $t_{i}$ is the $i$-th duration across all stages, and $n$ is the total number of duration entries across all stages.

- The summation is over all temporal durations $t_{i}$ across all stages.

3.8 **NREM Temporal Entropy**

The NREM Temporal Entropy is specifically calculated for the combined NREM stages (N1, N2, N3), measuring the randomness in the distribution of time durations spent within these stages.

$$H_{\text{NREM}\text{\_}\text{T}\text{emporal}}=-\sum_{k=1}^{l} p_{k}\log_{2} \left( p_{k} \right)$$

Where:

- $p_{k}$ is the probability associated with the $k$-th duration within the NREM stages.

- $p_{k}=\frac{t_{k}}{\sum_{j=1}^{l} t_{j}}$ , where $t_{k}$ is the $k$-th duration in NREM stages, and $l$ is the total number of duration entries for NREM stages.

- The summation is over all temporal durations $t_{k}$ spent in the NREM stages.

### 1 The Shanghai Sleep Health Study Cohort

#### 1.1 Correlation Heatmap in the SSHSC

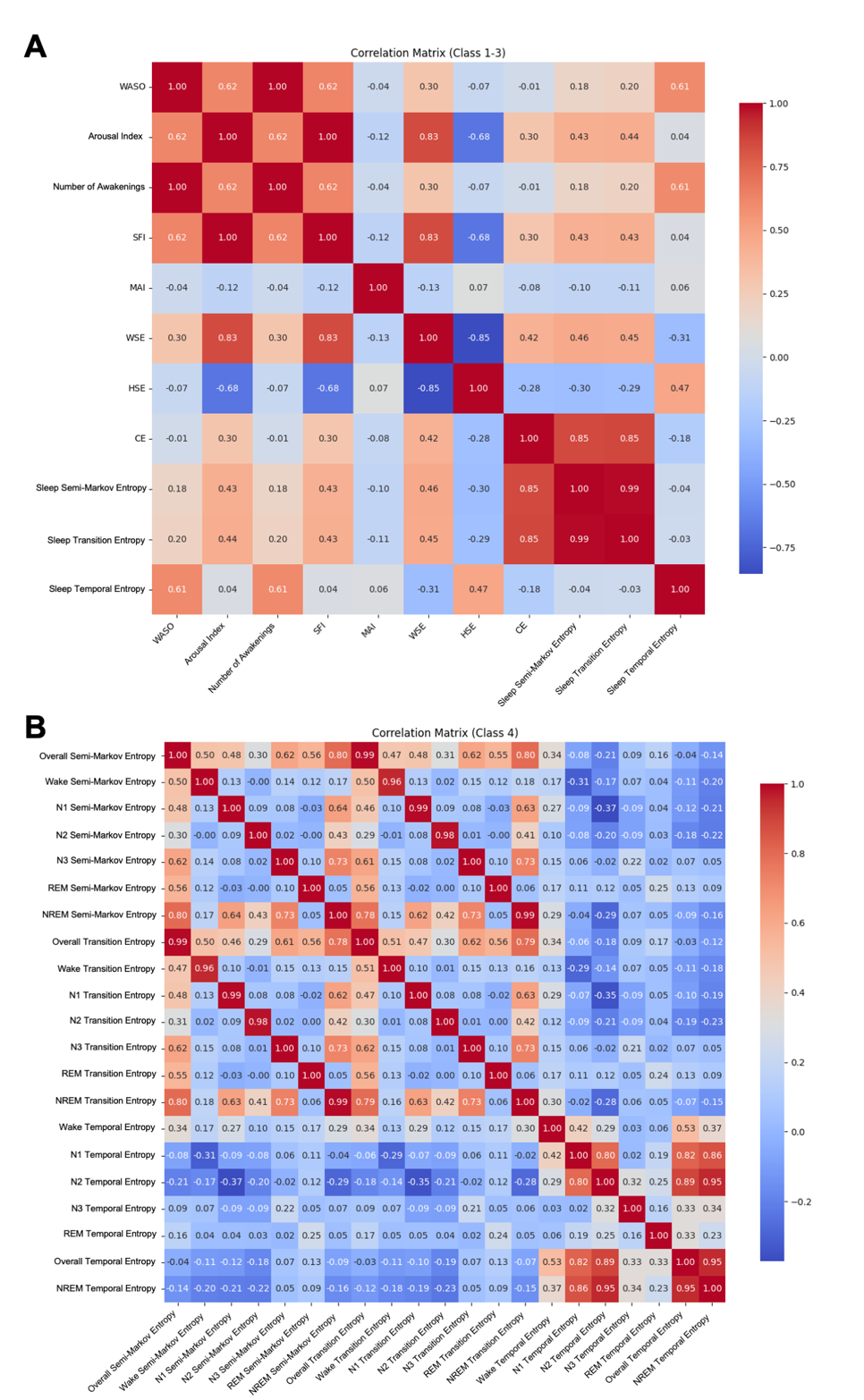

***Supplementary Figure 1: Correlation matrices of sleep fragmentation metrics categorized into different classes:*** ***Panel*** ***A*** *represents metrics from Class 1–3 in the SSHSC cohort: Class 1 includes traditional metrics such as WASO and Arousal Index (ArI); Class 2 consists of entropy-based metrics, including Haar Spectral Entropy (HSE), Walsh Spectral Entropy (WSE), and Conditional Entropy (CE); Class 3 introduces novel markers, such as Transition Entropy, Temporal Entropy, and Semi-Markov Entropy.* ***Panel B*** *extends the analysis to Class 4, including stage-specific entropy metrics (e.g., NREM, REM, N1, N2, and N3 STE), highlighting correlations across sleep stages and overall-level entropy. Correlation coefficients are color-coded, with red representing strong positive correlations and blue indicating strong negative correlations.*

#### 1.2 Post-hoc Explanation in Machine Learning in the SSHSC

| **Supplementary Table 1: Performance of XGBoost Model for Predicting Disease Outcomes** | | | | | |
| --- | --- | --- | --- | --- | --- |
|  | **Accuracy** | **Precision** | **Recall** | **F1** | **ROC AUC** |
| **Hypertension** | 0.737 | 0.521 | 0.295 | 0.375 | 0.697 |
| **Diabetes** | 0.810 | 0.295 | 0.500 | 0.369 | 0.754 |
| **Hyperlipidemia** | 0.630 | 0.606 | 0.720 | 0.658 | 0.669 |
| *All models used XGBoost (XGBClassifier) with binary: logistic as the objective function for binary classification tasks. Cross-validation: A 5-fold cross-validation strategy was employed, with each fold using training and validation to evaluate model performance, ensuring robust generalization.* | | | | | |

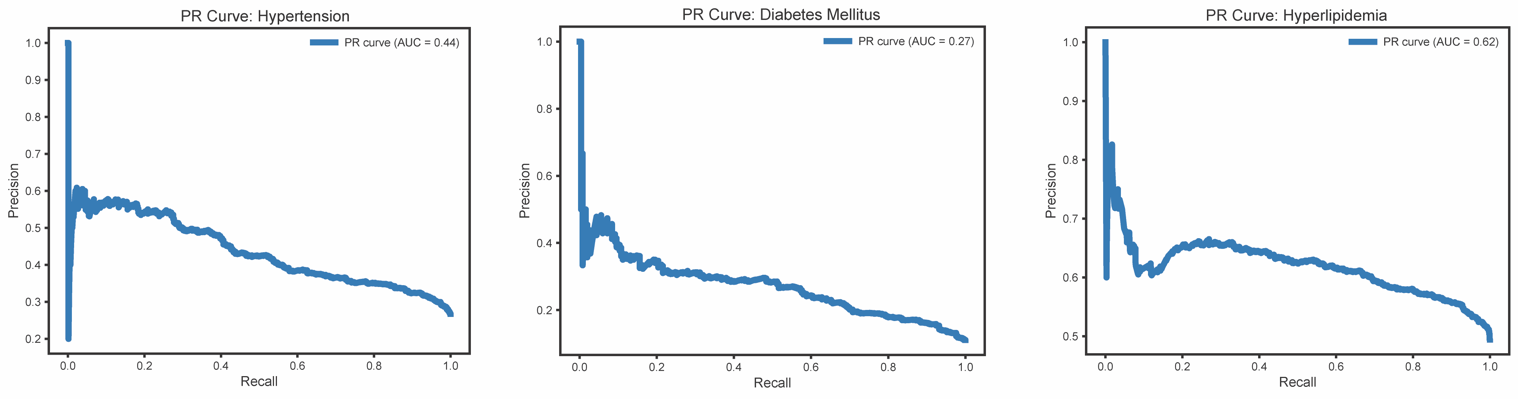

***Supplementary Figure 2: Precision–Recall Curves for Cardiometabolic Condition Prediction in the SSHSC:*** *Precision–recall (PR) curves for predicting cardiometabolic conditions in the SSHSC, including hypertension (HTN), diabetes mellitus (DM), and hyperlipidemia. Model performance is evaluated using the area under the precision–recall curve (AUPRC). The PR curves illustrate the trade-off between precision and recall across different classification thresholds for each condition. Given the varying prevalence of these outcomes, PR curves provide a more informative assessment of model performance under class imbalance.*

### 2 The Sleep Heart Health Study

| **Supplementary Table 2: Cohort Medical History of the Sleep Heart Health Study** | |
| --- | --- |
| **Medical History** | **N = 4862** |
| **Cardiovascular conditions** |  |
| Hypertension, n (%) | 1590 (32.7) |
| Angina, n (%) | 395 (8.1) |
| Coronary Angioplasty, n (%) | 157 (3.2) |
| CABG, n (%) | 185 (3.8) |
| HF, n (%) | 89 (1.8) |
| MI, n (%) | 323 (6.6) |
| Other Heart/Cardiac Surgery, n (%) | 125 (2.6) |
| **Other comorbidities** |  |
| Diabetes, n (%) | 354 (7.3) |
| Asthma, n (%) | 424 (8.7) |
| COPD, n (%) | 56 (1.2) |
| Chronic Bronchitis, n (%) | 272 (5.6) |
| Emphysema, n (%) | 115 (2.4) |
| Sleep Apnea, n (%) | 47 (1.0) |
| Sleep Medicine Use, n (%) |  |
| Never | 3244 (66.7) |
| Rarely | 425 (8.7) |
| Sometimes | 264 (5.4) |
| Often | 127 (2.6) |
| Almost always | 215 (4.4) |

*N: Number of subjects;* *Medical History: refers to the diseases that were self-reported by the patients during the first visit of the SHHS as having been diagnosed by a Doctor of Medicine. CABG: Coronary artery bypass graft; HF: Heart failure; MI: Myocardial infarction; COPD: Chronic obstructive pulmonary disease.*

#### 2.1 Correlation Heatmap in the SHHS

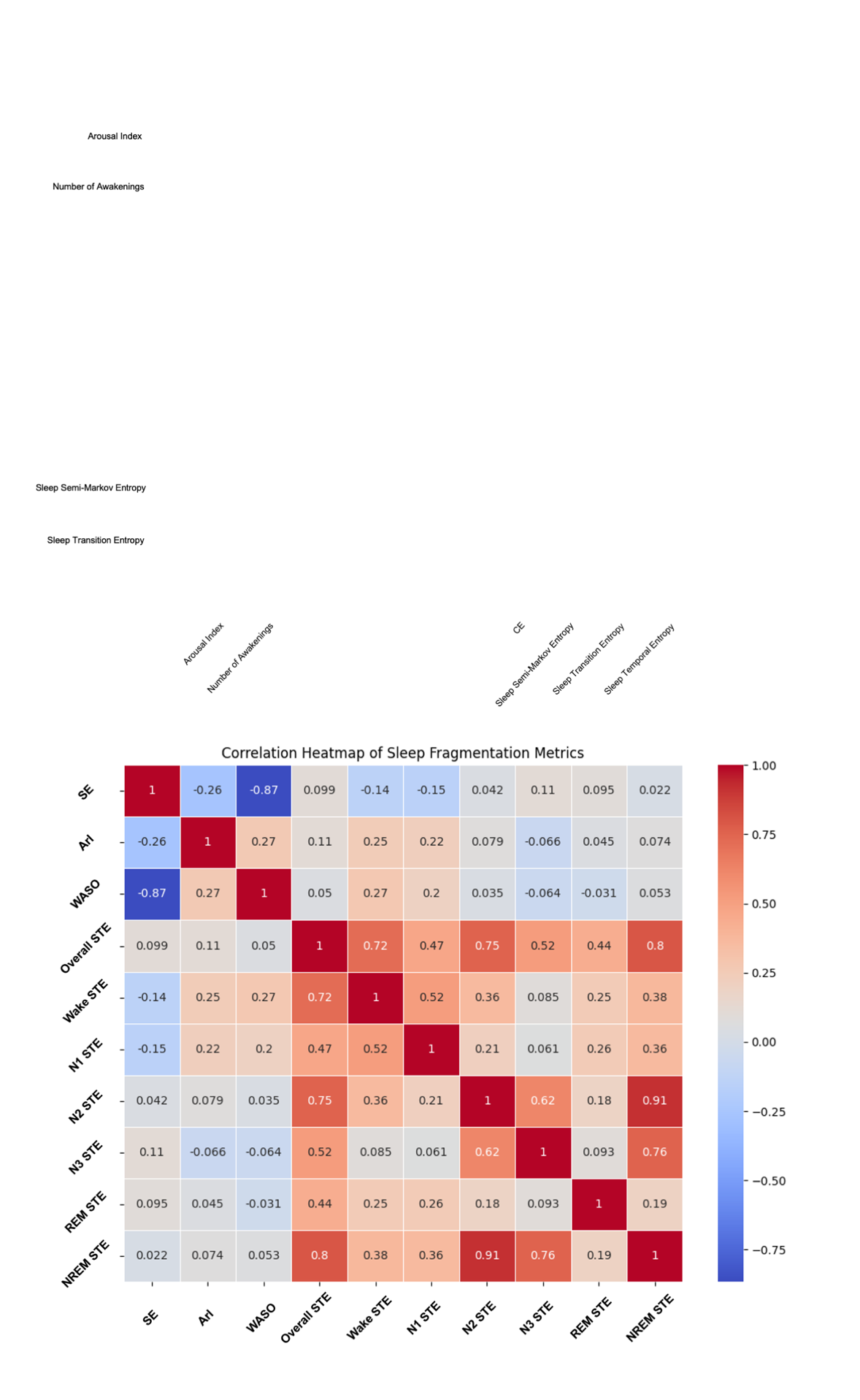

***Supplementary Figure 3: Correlation heatmap of sleep fragmentation metrics in the SHHS cohort:*** *Comparing traditional metrics (Sleep Efficiency [SE], Arousal Index [ArI], and WASO) with both whole-night and stage-specific Sleep Temporal Entropy (STE). This analysis builds on findings from the SSHSC, where STE demonstrated superior overall performance. The heatmap illustrates the relationships between traditional and entropy-based metrics, with red indicating positive correlations and blue representing negative correlations.*

#### 2.2 Post-hoc Explanation in Machine Learning in the SHHS

| **Supplementary Table 3: Performance of Different Machine Learning Models for Predicting All-Cause Mortality** | | | | | |
| --- | --- | --- | --- | --- | --- |
| **Model** | | **Accuracy** | **Precision** | **Recall** | **ROC AUC** |
| **XGBoost** | 5-Fold CV Mean | 0.861 | 0.670 | 0.243 | 0.812 |
|  | Test | 0.870 | 0.650 | 0.250 | 0.836 |
| **RF** | 5-Fold CV Mean | 0.838 | 0.489 | 0.498 | 0.818 |
|  | Test | 0.847 | 0.496 | 0.611 | 0.853 |
| **SVM** | 5-Fold CV Mean | 0.725 | 0.329 | 0.713 | 0.792 |
|  | Test | 0.864 | 0.634 | 0.246 | 0.826 |
| **KNN** | 5-Fold CV Mean | 0.828 | 0.421 | 0.232 | 0.667 |
|  | Test | 0.821 | 0.337 | 0.186 | 0.637 |
| **Logistic Regression** | 5-Fold CV Mean | 0.709 | 0.321 | 0.75 | 0.793 |
|  | Test | 0.724 | 0.331 | 0.805 | 0.829 |

*Performance metrics of different machine learning models for predicting all-cause mortality in the SHHS cohort. Metrics include accuracy, precision, recall, and ROC AUC, presented for both 5-fold cross-validation (CV) means and test datasets. Models evaluated include XGBoost, Random Forest (RF), Support Vector Machine (SVM), k-Nearest Neighbors (KNN), and Logistic Regression. ROC AUC reflects the area under the receiver operating characteristic curve, while precision and recall evaluate the balance between positive predictions and actual positive cases. XGBoost and RF demonstrated the highest ROC AUC values across both cross-validation and test datasets.*

| **Supplementary Table 4: Performance of Different Machine Learning Models for Predicting CVD Mortality** | | | | | |
| --- | --- | --- | --- | --- | --- |
| **Model** | | **Accuracy** | **Precision** | **Recall** | **ROC AUC** |
| **XGBoost** | 5-Fold CV Mean | 0.952 | 0.326 | 0.036 | 0.864 |
|  | Test | 0.952 | 0.600 | 0.075 | 0.838 |
| **RF** | 5-Fold CV Mean | 0.937 | 0.345 | 0.272 | 0.858 |
|  | Test | 0.934 | 0.34 | 0.375 | 0.796 |
| **SVM** | 5-Fold CV Mean | 0.772 | 0.144 | 0.774 | 0.846 |
|  | Test | 0.951 | 0.500 | 0.025 | 0.815 |
| **KNN** | 5-Fold CV Mean | 0.941 | 0.109 | 0.026 | 0.591 |
|  | Test | 0.939 | 0 | 0 | 0.545 |
| **Logistic Regression** | 5-Fold CV Mean | 0.765 | 0.149 | 0.838 | 0.849 |
|  | Test | 0.759 | 0.134 | 0.725 | 0.804 |

*Performance metrics of different machine learning models for predicting CVD mortality in the SHHS cohort. Metrics include accuracy, precision, recall, and ROC AUC, presented for both 5-fold cross-validation (CV) means and test datasets. Models evaluated include XGBoost, Random Forest (RF), Support Vector Machine (SVM), k-Nearest Neighbors (KNN), and Logistic Regression. ROC AUC reflects the area under the receiver operating characteristic curve, while precision and recall evaluate the balance between positive predictions and actual positive cases. XGBoost and RF demonstrated the highest ROC AUC values across both cross-validation and test datasets.*

#### 2.3 Model Evaluation and Comparison in the SHHS

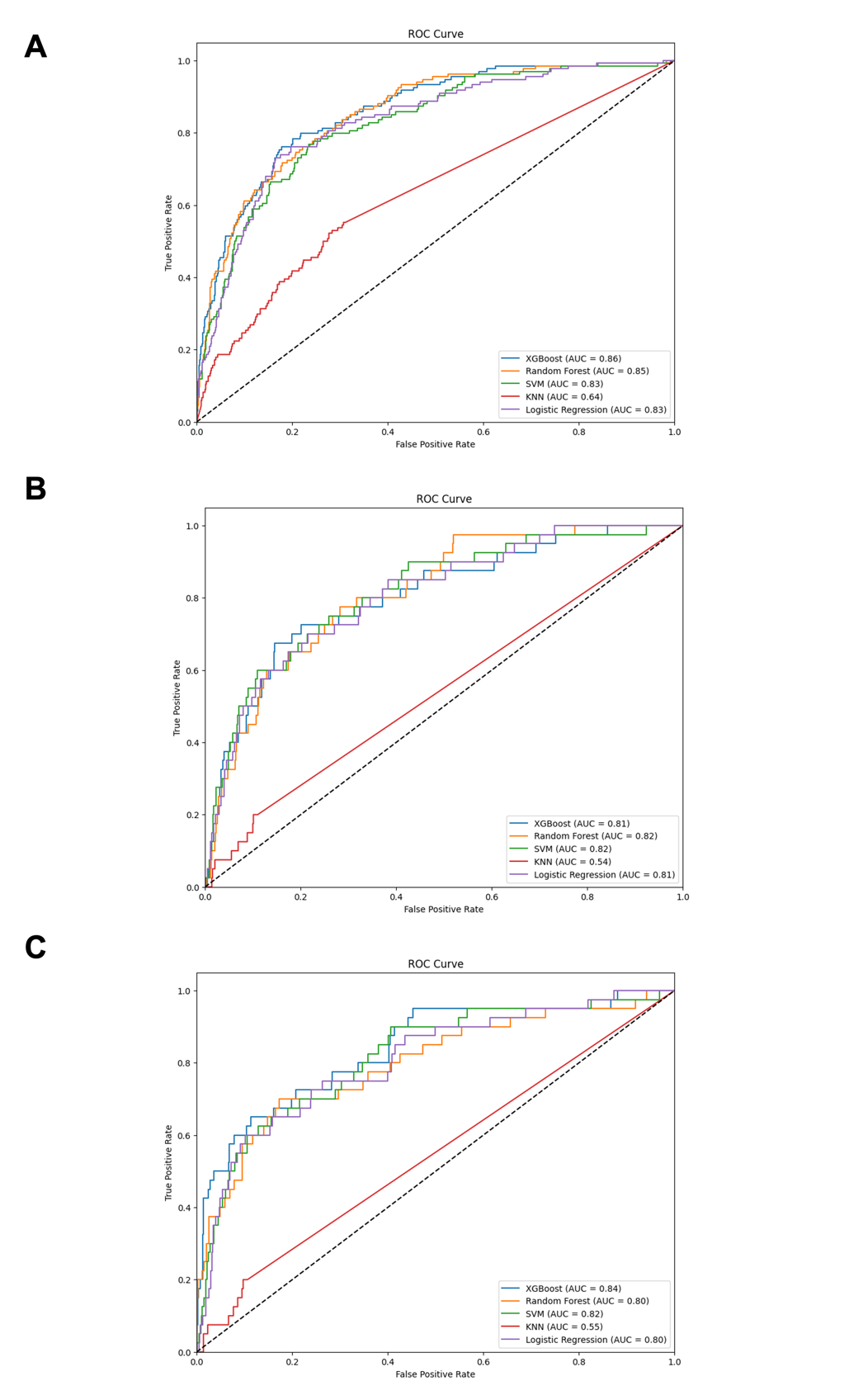

***Supplementary Figure 4: Panels A–C represent the Receiver Operating Characteristic (ROC) curves for different models****: (****A****) Whole-Night Metrics for All-Cause Mortality, (****B****) Whole-Night Metrics for CVD Mortality, and (****C****) Stage-Specific Metrics for CVD Mortality. Each panel compares the performance of various machine learning models, including XGBoost, Random Forest, SVM, KNN, and Logistic Regression, as indicated by their respective AUC values.*

| **Supplementary Table 5: Comparison of Model Performance Metrics with and without STE** | | | | | |
| --- | --- | --- | --- | --- | --- |
| **Input** | **Accuracy** | **Precision** | **Recall** | **F1 Score** | **ROC AUC Score** |
| **without markers** | 0.86532 | 0.627119 | 0.274074 | 0.381443 | 0.823682 |
| **with markers** | 0.874718 | 0.709091 | 0.291045 | 0.412698 | 0.85801 |
| **Difference Value** | 0.009398 | 0.081972 | 0.016971 | 0.031255 | 0.034328 |
| *The results presented in this table are based on the all-cause mortality data from the Sleep Heart Health Study (SHHS). ROC AUC (Receiver Operating Characteristic Area Under the Curve) is used to evaluate the classification performance of the models. Accuracy refers to the proportion of correctly classified samples, while precision is the proportion of true positive predictions out of all positive predictions. Recall measures the proportion of actual positive samples correctly predicted by the model, and F1 Score is the harmonic mean of precision and recall, providing a balanced measure of model performance. All-cause mortality represents the risk of death from all causes. The Difference column shows the change in model performance when Sleep Temporal Entropy (STE) is added, indicating whether the inclusion of STE improves the predictive capability of the model.* | | | | | |

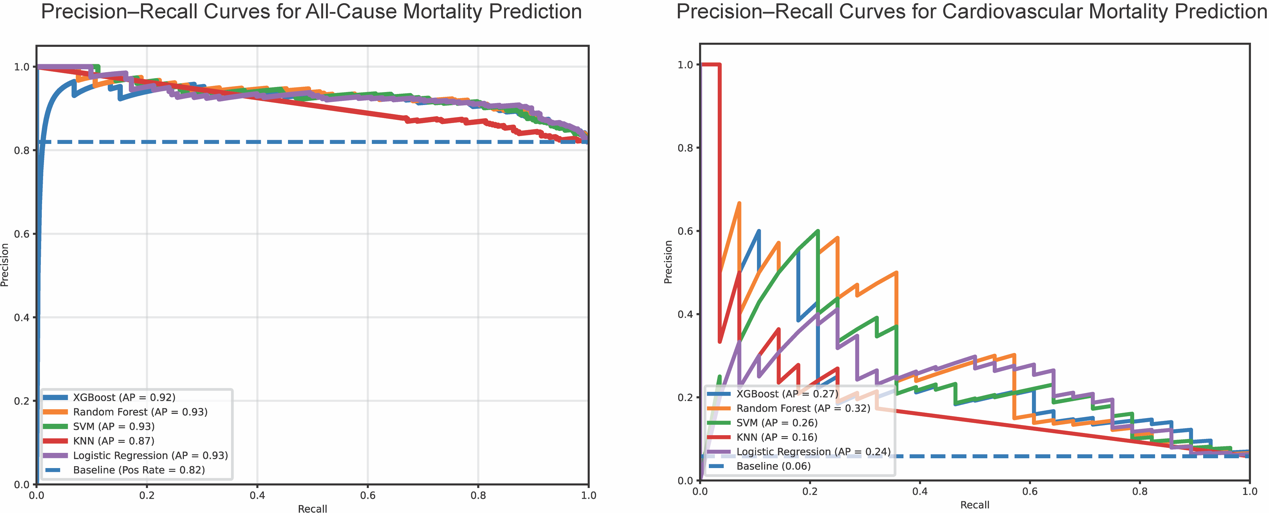

***Supplementary Figure 5: Precision–recall (PR) curves for predicting all-cause mortality and cardiovascular (CVD) mortality in the SHHS cohort：*** *Model performance is summarized by the area under the precision–recall curve (AUPRC). The PR curves demonstrate the trade-off between precision and recall across classification thresholds for multiple machine learning models. The horizontal dashed line in each panel represents the outcome prevalence in the test set. Notably, the higher baseline in all-cause mortality compared to CVD mortality indicates differing degrees of class imbalance, which is appropriately captured by PR-based evaluation.*

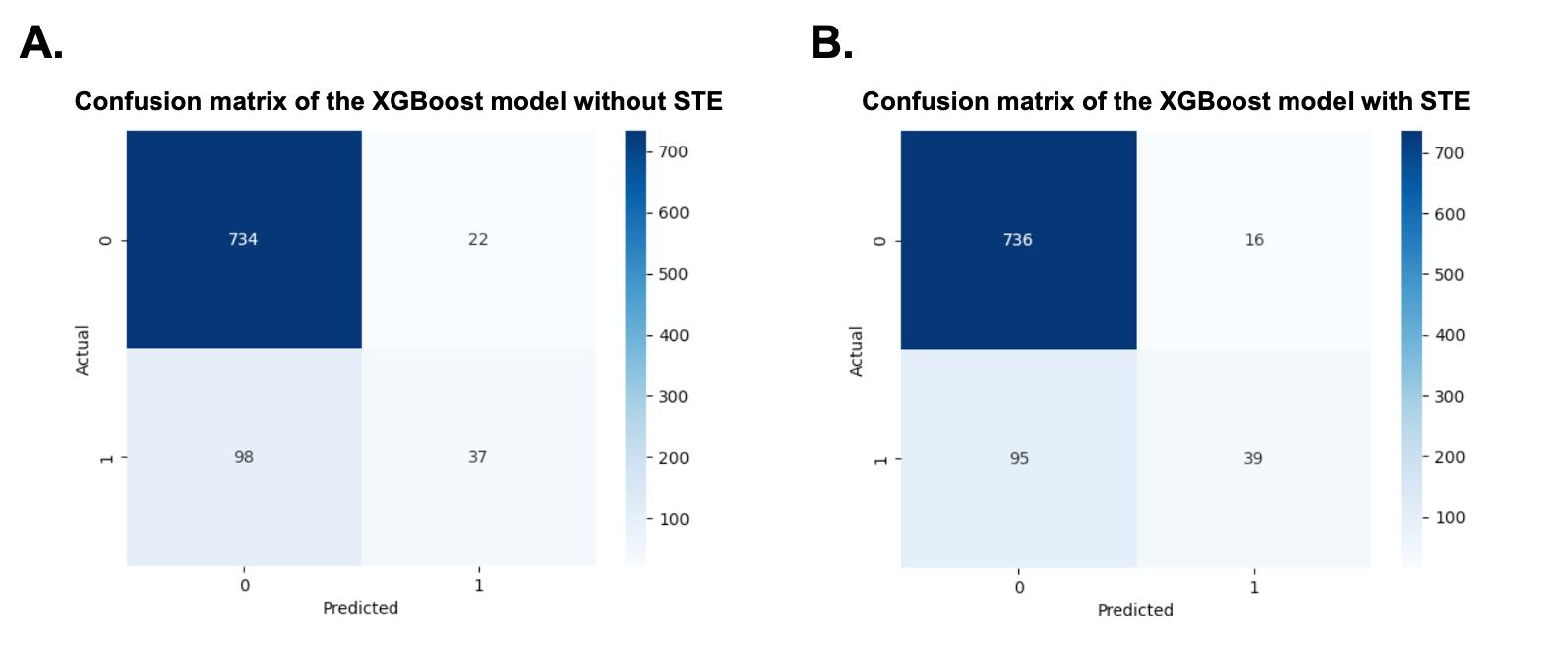

***Supplementary Figure 6: Confusion matrices for all-cause mortality prediction using the XGBoost model, with and without the inclusion of Sleep Temporal Entropy (STE). Panel A*** *shows the confusion matrix for the model without STE, and* ***Panel B*** *presents the results after incorporating STE. The inclusion of STE slightly improves classification performance by increasing the number of true positives (from 37 to 39) and reducing false positives (from 22 to 16), suggesting modest but measurable enhancement in model sensitivity and precision.*

#### 2.4 Survival Analysis

**2.4.1 Cohort Characteristics and Proportional Hazards Assumption Test**

| **Supplementary Table 6: Cohort characteristics of the Sleep Heart Health Study by All-cause and CVD Mortality** | | | |
| --- | --- | --- | --- |
| **Characteristics** | **Alive**  **(n = 3743)** | **All-cause mortality**  **(n = 1119)** | **CVD Mortality**  **(n = 344)** |
| **Basic characteristics** |  |  |  |
| Age (years), median (IQR) | 60.0 [54.0, 69.0] | 75.0 [69.0, 79.5] | 76.0 [72.8, 81.0] |
| Gender, n(%) |  |  |  |
| Male | 1657 (44.3) | 589 (52.6) | 188 (54.7) |
| Female | 2086 (55.7) | 530 (47.4) | 156 (45.3) |
| Race, n(%) |  |  |  |
| White | 3244 (86.7) | 981 (87.7) | 295 (85.8) |
| Black | 204 (5.5) | 113 (10.1) | 47 (13.7) |
| Other | 295 (7.9) | 25 (2.2) | 2 (0.6) |
| BMI (kg/m2), median (IQR) | 27.8 [24.9, 30.9] | 27.0 [24.3, 30.5] | 27.1 [24.1, 30.1] |
| Smoking, n(%) | 361 (9.6) | 107 (9.6) | 23 (6.7) |
| **Medical History** |  |  |  |
| Hypertension, n(%) | 1041 (27.8) | 549 (49.1) | 197 (57.3) |
| Angina, n(%) | 226 (6.0) | 169 (15.1) | 69 (20.1) |
| Coronary Angioplasty, n(%) | 92 (2.5) | 65 (5.8) | 25 (7.3) |
| CABG, n(%) | 82 (2.2) | 103 (9.2) | 45 (13.1) |
| HF, n(%) | 27 (0.7) | 62 (5.5) | 24 (7.0) |
| MI, n(%) | 160 (4.3) | 163 (14.6) | 74 (21.5) |
| Other Heart/Cardiac Surgery, n(%) | 68 (1.8) | 57 (5.1) | 27 (7.8) |
| Diabetes, n(%) | 179 (4.8) | 175 (15.6) | 78 (22.7) |
| Asthma, n(%) | 336 (9.0) | 88 (7.9) | 28 (8.1) |
| COPD, n(%) | 31 (0.8) | 25 (2.2) | 4 (1.2) |
| Chronic Bronchitis, n(%) | 186 (5.0) | 86 (7.7) | 20 (5.8) |
| Emphysema, n(%) | 57 (1.5) | 58 (5.2) | 19 (5.5) |
| Sleep Apnea, n(%) | 33 (0.9) | 14 (1.3) | 2 (0.6) |
| Sleeppills, n(%) |  |  |  |
| Never | 2568 (68.6) | 676 (60.4) | 218 (63.4) |
| Rarely | 339 (9.1) | 86 (7.7) | 22 (6.4) |
| Sometimes | 207 (5.5) | 57 (5.1) | 15 (4.4) |
| Often | 93 (2.5) | 34 (3.0) | 8 (2.3) |
| Almost always | 158 (4.2) | 57 (5.1) | 22 (6.4) |
| ***Note*** *n: Number of subjects; IQR: Interquartile range; BMI: Body mass index; CABG: Coronary artery bypass graft; HF: Heart failure; MI: Myocardial infarction; COPD: Chronic obstructive pulmonary disease; WASO: Wake after sleep onset.* | | | |

| **Supplementary Table 7: Cohort characteristics of the Sleep Heart Health Study by All-cause and CVD Mortality** | | | |
| --- | --- | --- | --- |
| **Characteristics** | **Alive**  **(n = 3743)** | **All-cause mortality**  **(n = 1119)** | **CVD Mortality**  **(n = 344)** |
| **PSG** |  |  |  |
| Total Sleep Time(mins), median (IQR) | 376.5 [332.0, 413.5] | 355.0 [306.8, 390.5] | 359.0 [297.3, 395.0] |
| N1 Sleep Time(mins), median (IQR) | 16.5 [10.0, 25.0] | 17.5 [10.5, 28.5] | 17.5 [10.0, 25.0] |
| N2 Sleep Time(mins), median (IQR) | 207.5 [170.5, 240.5] | 201.0 [160.0, 239.0] | 204.0 [154.5, 249.0] |
| N3 Sleep Time(mins), median (IQR) | 65.5 [36.0, 94.0] | 54.5 [24.0, 89.5] | 52.0 [20.5, 86.0] |
| REM Sleep Time(mins), median (IQR) | 76.5 [58.0, 94.5] | 64.0 [45.5, 82.0] | 62.0 [45.5, 83.0] |
| Sleep Efficiency (%), median (IQR) | 86.2 [79.4, 91.0] | 81.1 [73.1, 88.0] | 81.1 [73.7, 87.1] |
| Arousal Index(events/hour), median (IQR) | 16.4 [11.8, 22.9] | 18.2 [12.6, 25.7] | 18.2 [13.0, 25.7] |
| WASO (minutes), median (IQR) | 46.5 [29.0, 75.5] | 66.5 [38.0, 103.8] | 70.8 [44.9, 105.1] |
| AHI (events/hour), median (IQR) | 12.3 [6.2, 21.6] | 14.6 [8.3, 24.1] | 16.1 [8.7, 24.7] |
| Hypoxic Burden (%·min/h), median (IQR) | 38.3 [19.8, 68.9] | 51.9 [26.8, 86.9] | 54.6 [32.1, 94.8] |
| T90 Category (%), n (%) |  |  |  |
| 0 | 3111 (83.1) | 883 (78.9) | 278 (80.8) |
| 0-5 | 25 (0.7) | 10 (0.9) | 0 (0.0) |
| >5 | 607 (16.2) | 226 (20.2) | 66 (19.2) |
| **Sleep Temporal Entropy (STE)** |  |  |  |
| Wake STE, median (IQR) | 2.6 [2.0, 3.2] | 2.7 [2.1, 3.3] | 2.8 [2.2, 3.4] |
| N1 STE, median (IQR) | 3.8 [3.3, 4.3] | 3.9 [3.3, 4.4] | 3.9 [3.3, 4.4] |
| N2 STE, median (IQR) | 4.5 [4.2, 4.9] | 4.5 [4.0, 4.9] | 4.5 [4.1, 4.8] |
| N3 STE, median (IQR) | 3.5 [2.9, 4.0] | 3.5 [2.8, 4.0] | 3.4 [2.7, 4.0] |
| REM STE, median (IQR) | 2.6 [2.1, 3.0] | 2.4 [1.8, 2.9] | 2.3 [1.9, 3.0] |
| Overall STE, median (IQR) | 5.5 [5.1, 5.8] | 5.4 [4.9, 5.8] | 5.4 [4.9, 5.8] |
| NREM STE, median (IQR) | 5.2 [4.8, 5.6] | 5.2 [4.7, 5.6] | 5.2 [4.7, 5.6] |
| ***Note*** *n: Number of subjects; IQR: Interquartile range; T90: proportion of total sleep time with oxygen saturation < 90%. Participants were categorized into 0%, 0–5%, and >5% groups; STE: Sleep temporal entropy.* | | | |

| **Supplementary Table 8: Proportional Hazards Assumption Test Results for Mortality Outcomes in the Sleep Heart Health Study** | | |
| --- | --- | --- |
| **STE** | **P-value**  **(All-Cause Mortality)** | **P-value**  **(CVD Mortality)** |
| **Overall STE** | 0.400 | 0.280 |
| **Wake STE** | 0.450 | 0.760 |
| **N1 STE** | 0.0007 | 0.770 |
| **N2 STE** | 0.0008 | 0.012 |
| **N3 STE** | 0.074 | 0.860 |
| **REM STE** | 0.490 | 0.250 |
| **NREM STE** | 0.001 | 0.001 |
| *Note: The P values presented above were derived from tests of the proportional hazard assumption using Schoenfeld residuals. P values were calculated using two-sided tests, with no adjustment for multiple comparisons. A P value < 0.05 indicates a violation of the proportional hazard assumption, suggesting that the effect of the covariate may vary over time. Variables were categorized into quintiles for analysis.* | | |

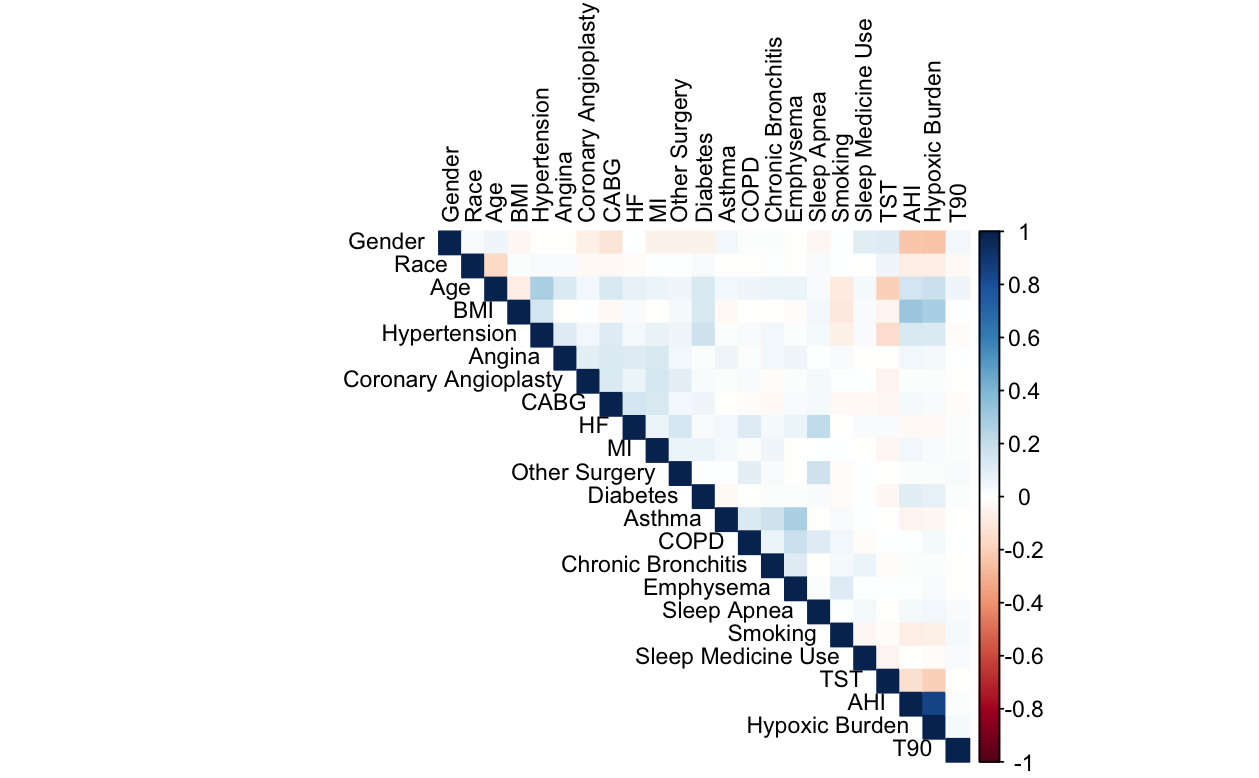

***Supplementary Figure 7: Heatmap of Pearson correlation coefficients among covariates included in the multivariable models:*** *Blue indicates positive correlations, and red indicates negative correlations. Darker colors represent stronger correlations.*

*Abbreviations: BMI: Body mass index; CABG: Coronary artery bypass graft; HF: Heart failure; MI: Myocardial infarction; COPD: Chronic obstructive pulmonary disease; TST: Total sleep time; AHI: Apnea–hypopnea index; T90: Proportion of total sleep time with oxygen saturation <90%; Hypoxic Burden: Integrated measure of the area under the desaturation curve during apneic/hypopneic events.*

**2.4.2 Sensitivity Analyses of Sleep Temporal Entropy Metrics and Mortality Outcomes**

| **Supplementary Table 9: Variance Inflation Factors (VIF) for Covariates in CVD and All-Cause Mortality Models** | | |
| --- | --- | --- |
| **Variable** | **VIF** | |
|  | **CVD Mortality** | **All-Cause Mortality** |
| Gender | 1.14 | 1.14 |
| Race | 1.05 | 1.05 |
| Age | 1.29 | 1.29 |
| BMI | 1.21 | 1.21 |
| Hypertension | 1.17 | 1.17 |
| Angina | 1.07 | 1.07 |
| Coronary Angioplasty | 1.05 | 1.05 |
| CABG | 1.10 | 1.10 |
| HF | 1.12 | 1.12 |
| MI | 1.06 | 1.06 |
| Other Heart/Cardiac Surgery | 1.07 | 1.07 |
| Diabetes | 1.07 | 1.07 |
| Asthma | 1.12 | 1.12 |
| COPD | 1.08 | 1.08 |
| Chronic Bronchitis | 1.06 | 1.06 |
| Emphysema | 1.14 | 1.14 |
| Sleep Apnea | 1.09 | 1.09 |
| Smoking | 1.05 | 1.05 |
| Sleep Medicine Use | 1.03 | 1.03 |
| TST | 1.11 | 1.11 |
| AHI | 3.61 | 3.61 |
| Hypoxic Burden | 3.59 | 3.59 |
| T90 | 1.01 | 1.01 |
| ***Note:*** *Variance inflation factors (VIFs) were calculated to assess multicollinearity among covariates included in the regression models. VIF values greater than 5 were considered indicative of potential multicollinearity. BMI, body mass index; CABG, coronary artery bypass graft; HF, heart failure; MI, myocardial infarction; COPD, chronic obstructive pulmonary disease; TST, total sleep time; AHI, apnea–hypopnea index; T90, proportion of total sleep time with oxygen saturation <90%; Hypoxic burden, integrated measure of the area under the desaturation curve during apneic/hypopneic events.* | | |

| **Supplementary Table 10: Sensitivity Analysis of Sleep Temporal Entropy and CVD Mortality Risk Across Quintiles** | | | | | |
| --- | --- | --- | --- | --- | --- |
| **Fragmentation Metric** | **HR (95% CI)** | | | | |
|  | **Q1** | **Q2** | **Q3** | **Q4** | **Q5** |
| Overall STE | 0.53 ( 0.26 - 1.08 ) | 0.94 ( 0.54 - 1.63 ) | 1 [Reference] | 0.90 ( 0.52 - 1.56 ) | 0.60 ( 0.34 - 1.04 ) |
| Wake STE | 0.83 ( 0.44 - 1.57 ) | 0.70 ( 0.36 - 1.33 ) | 1 [Reference] | 1.41 ( 0.82 - 2.41 ) | 1.14 ( 0.65 - 1.99 ) |
| REM STE | 1.65 ( 0.88 - 3.11 ) | 1.37 ( 0.70 - 2.69 ) | 1 [Reference] | 1.76 ( 0.90 - 3.47 ) | **2.40 ( 1.27 - 4.52 )** |
| N1 STE | 1.24 ( 0.68 - 2.26 ) | 0.97 ( 0.52 - 1.79 ) | 1 [Reference] | 1.27 ( 0.73 - 2.23 ) | 1.17 ( 0.66 - 2.09 ) |
| N2 STE | 2.22 ( 1.03 - 4.80 ) | 2.70 ( 1.33 - 5.48 ) | 1 [Reference] | 2.31 ( 1.17 - 4.56 ) | 1.60 ( 0.78 - 3.30 ) |
| N3 STE | 0.83 ( 0.46 - 1.50 ) | 0.85 ( 0.51 - 1.42 ) | 1 [Reference] | 0.75 ( 0.44 - 1.26 ) | **0.39 ( 0.21 - 0.71 )** |
| NREM STE | 0.94 ( 0.49 - 1.79 ) | 1.13 ( 0.65 - 1.98 ) | 1 [Reference] | 0.77 ( 0.46 - 1.31 ) | **0.53 ( 0.30 - 0.93 )** |
| *STE, Sleep Temporal Entropy. Hazard ratios (HRs) and corresponding 95% confidence intervals (CIs) were estimated using Cox proportional hazards models adjusting for gender, race, age, BMI, hypertension, diabetes, asthma, COPD, sleep apnea, smoking, sleep medication use, Apnea–Hypopnea Index (AHI), total sleep time (TST), hypoxic burden, and T90. P values were calculated using two-sided tests, with no adjustment for multiple comparisons. Bold values indicate statistically significant associations (REM STE Q5: P = 0.004; N3 STE Q2: P = 0.008; NREM STE Q5: P = 0.038).* | | | | | |

| **Supplementary Table 11: Sensitivity Analysis of Sleep Temporal Entropy and All-Cause Mortality Risk Across Quintiles** | | | | | |
| --- | --- | --- | --- | --- | --- |
| **Fragmentation Metric** | **HR (95% CI)** | | | | |
|  | **Q1** | **Q2** | **Q3** | **Q4** | **Q5** |
| Overall STE | 0.91 ( 0.76 - 1.09 ) | 0.91 ( 0.79 - 1.05 ) | 1 [Reference] | 1.02 ( 0.89 - 1.16 ) | 0.99 ( 0.86 - 1.13 ) |
| Wake STE | 0.96 ( 0.83 - 1.11 ) | 1.01 ( 0.88 - 1.16 ) | 1 [Reference] | 1.00 ( 0.87 - 1.15 ) | 1.03 ( 0.90 - 1.19 ) |
| REM STE | **0.79 ( 0.68 - 0.93 )** | 0.96 ( 0.84 - 1.11 ) | 1 [Reference] | 1.02 ( 0.90 - 1.17 ) | 0.98 ( 0.85 - 1.12 ) |
| N1 STE | 1.01 ( 0.88 - 1.17 ) | 0.97 ( 0.84 - 1.11 ) | 1 [Reference] | 1.01 ( 0.88 - 1.16 ) | 1.03 ( 0.89 - 1.19 ) |
| N2 STE | 0.84 ( 0.71 - 1.00 ) | 0.90 ( 0.78 - 1.04 ) | 1 [Reference] | 0.89 ( 0.78 - 1.01 ) | 0.98 ( 0.86 - 1.12 ) |
| N3 STE | 0.98 ( 0.84 - 1.13 ) | 0.99 ( 0.86 - 1.14 ) | 1 [Reference] | 1.02 ( 0.89 - 1.17 ) | 1.04 ( 0.90 - 1.19 ) |
| NREM STE | **0.83 ( 0.69 - 0.99 )** | 0.92 ( 0.80 - 1.06 ) | 1 [Reference] | 0.91 ( 0.80 - 1.04 ) | 0.94 ( 0.83 - 1.08 ) |
| *STE, Sleep Temporal Entropy. Hazard ratios (HRs) and corresponding 95% confidence intervals (CIs) were estimated using Cox proportional hazards models adjusting for gender, race, age, BMI, hypertension, diabetes, asthma, COPD, sleep apnea, smoking, sleep medication use, Apnea–Hypopnea Index (AHI), total sleep time (TST), hypoxic burden, and T90. P values were calculated using two-sided tests, with no adjustment for multiple comparisons. Bold values indicate statistically significant associations (REM STE Q1: P = 0.005; NREM STE Q1: P = 0.048).* | | | | | |

**2.4.3 Whole-Night Kaplan-Meier Survival Curves for Fully Adjusted Models**

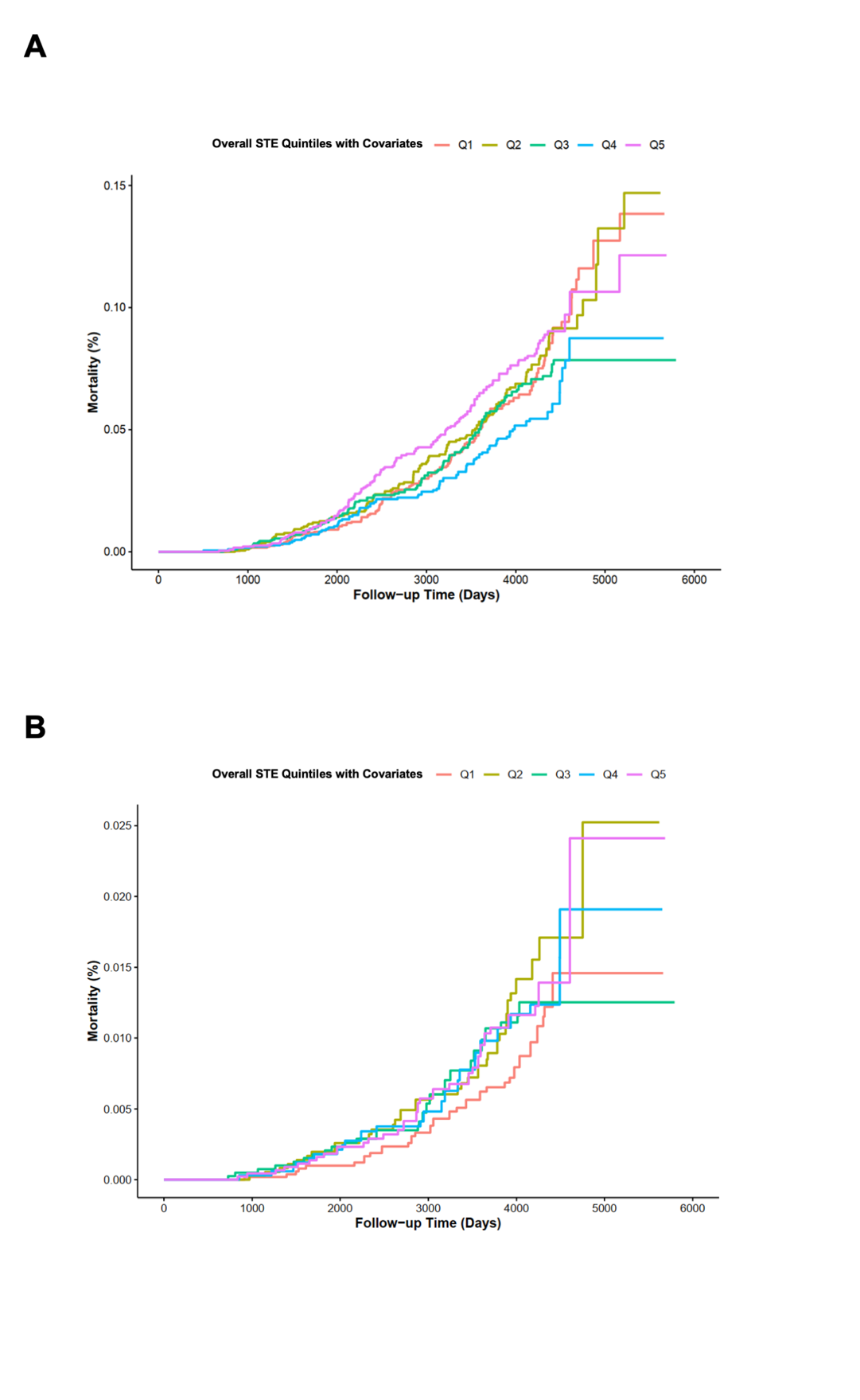

***Supplementary Figure 8: Panels A and B represent Kaplan-Meier (KM) survival curves for fully adjusted models****: (****A****) Whole-Night Metrics for All-Cause Mortality (Section 2.4.1) and (****B****) Whole-Night Metrics for CVD Mortality (Section 2.4.2). The fully adjusted models include covariates for demographic factors (gender, race, age, and BMI), comorbidities (hypertension, diabetes, asthma, COPD, and sleep apnea), lifestyle factors (smoking and sleep medication use), and total sleep time (TST). For stage-specific STE models (e.g., REM STE, N3 STE), the corresponding sleep stage duration (e.g., REM sleep time, N3 sleep time) is also included as a covariate.*

**2.4.3 Stage-Specific Kaplan-Meier Survival Curves All-Cause Mortality**

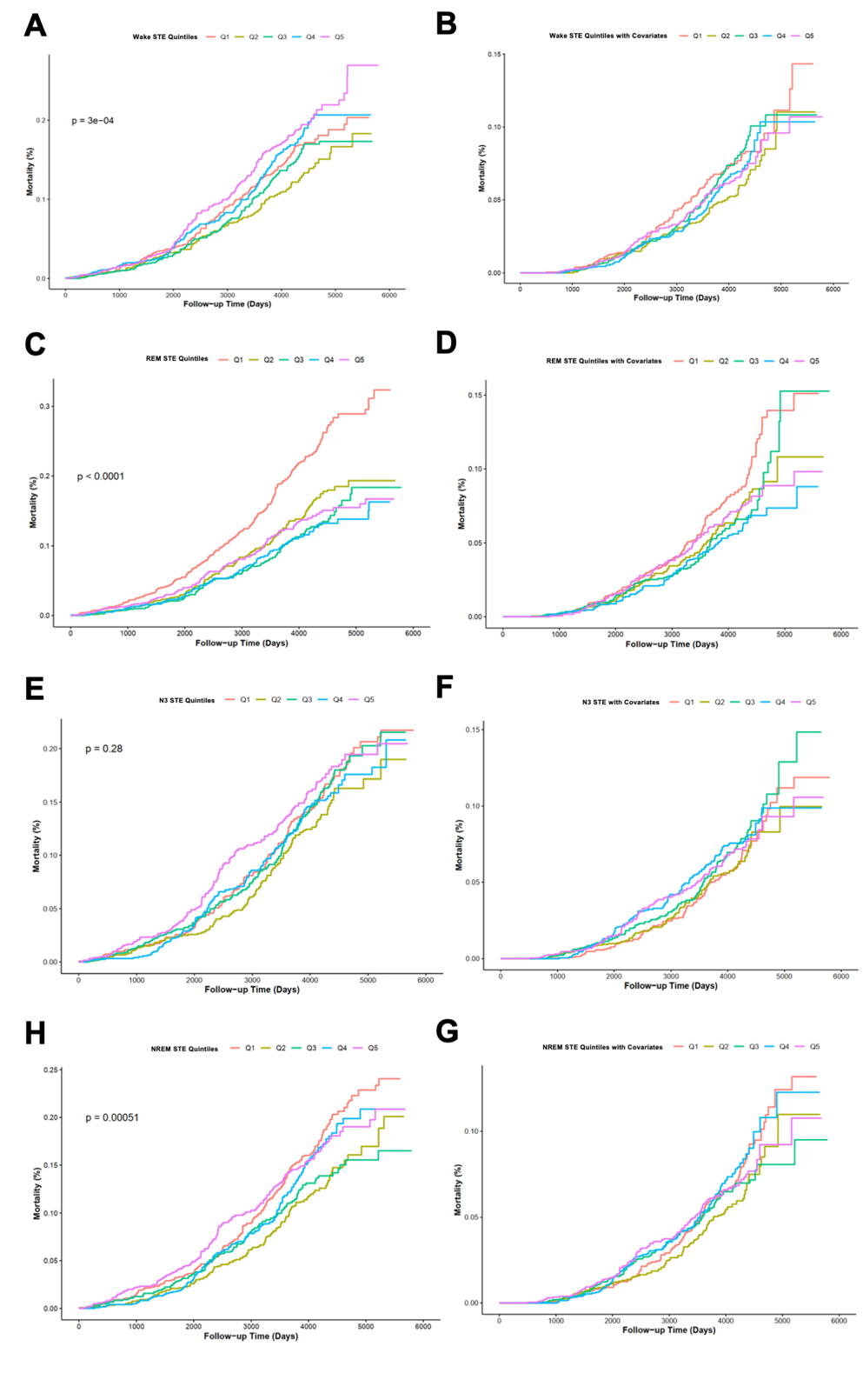

***Supplementary Figure 9: Kaplan-Meier (KM) curves for stage-specific sleep temporal entropy (STE) quintiles and all-cause mortality:*** *Panels A and B represent Wake STE, with (****A****) unadjusted and (****B****) fully adjusted models. Panels C and D display REM STE, with (****C****) unadjusted and (****D****) fully adjusted models. Panels E and F show N3 STE, with (****E****) unadjusted and (****F****) fully adjusted models. Panels G and H depict NREM STE, with (****G****) unadjusted and (****H****) fully adjusted models. Fully adjusted models account for demographic factors (gender, race, age, BMI), comorbidities (hypertension, diabetes, asthma, COPD, sleep apnea), lifestyle factors (smoking, sleep medication use), and total sleep time (TST). For stage-specific STE, the corresponding sleep stage duration (e.g., REM sleep time, N3 sleep time) is also included as a covariate.*

**2.4.3 Stage-Specific Kaplan-Meier Survival Curves for CVD Mortality**

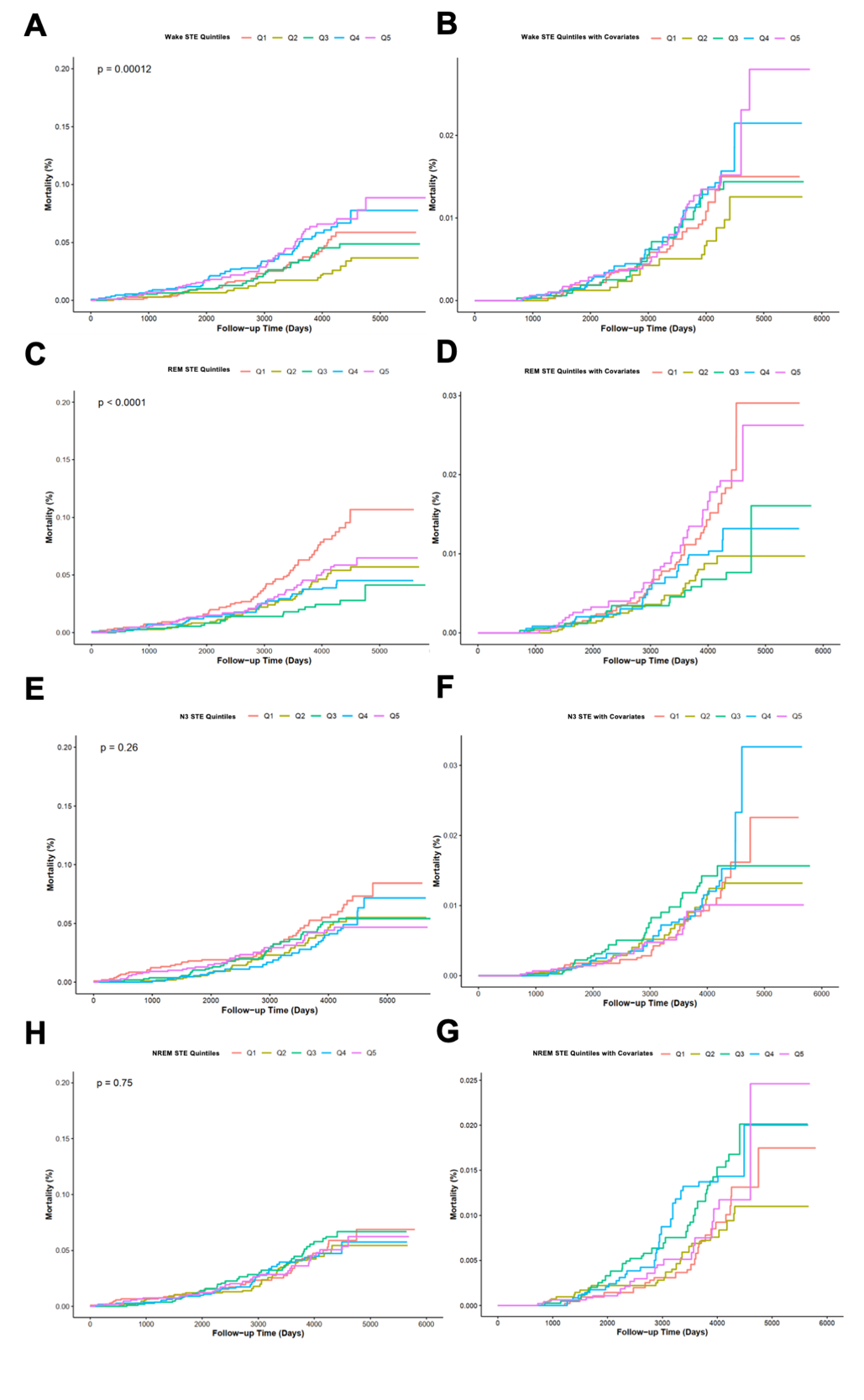

***Supplementary Figure 10: Kaplan-Meier (KM) curves for stage-specific sleep temporal entropy (STE) quintiles and CVD mortality****: Panels A and B represent Wake STE, with (****A****) unadjusted and (****B****) fully adjusted models. Panels C and D display REM STE, with (****C****) unadjusted and (****D****) fully adjusted models. Panels E and F show NREM STE, with (****E****) unadjusted and (****F****) fully adjusted models. Panels G and H depict N3 STE, with (****G****) unadjusted and (****H****) fully adjusted models.* *Fully adjusted models include the same covariates as described previously.*

#### 2.5 Restricted Cubic Spline

**2.5.1 Stage-Specific Restricted Cubic Spline Curves for All-Cause Mortality**

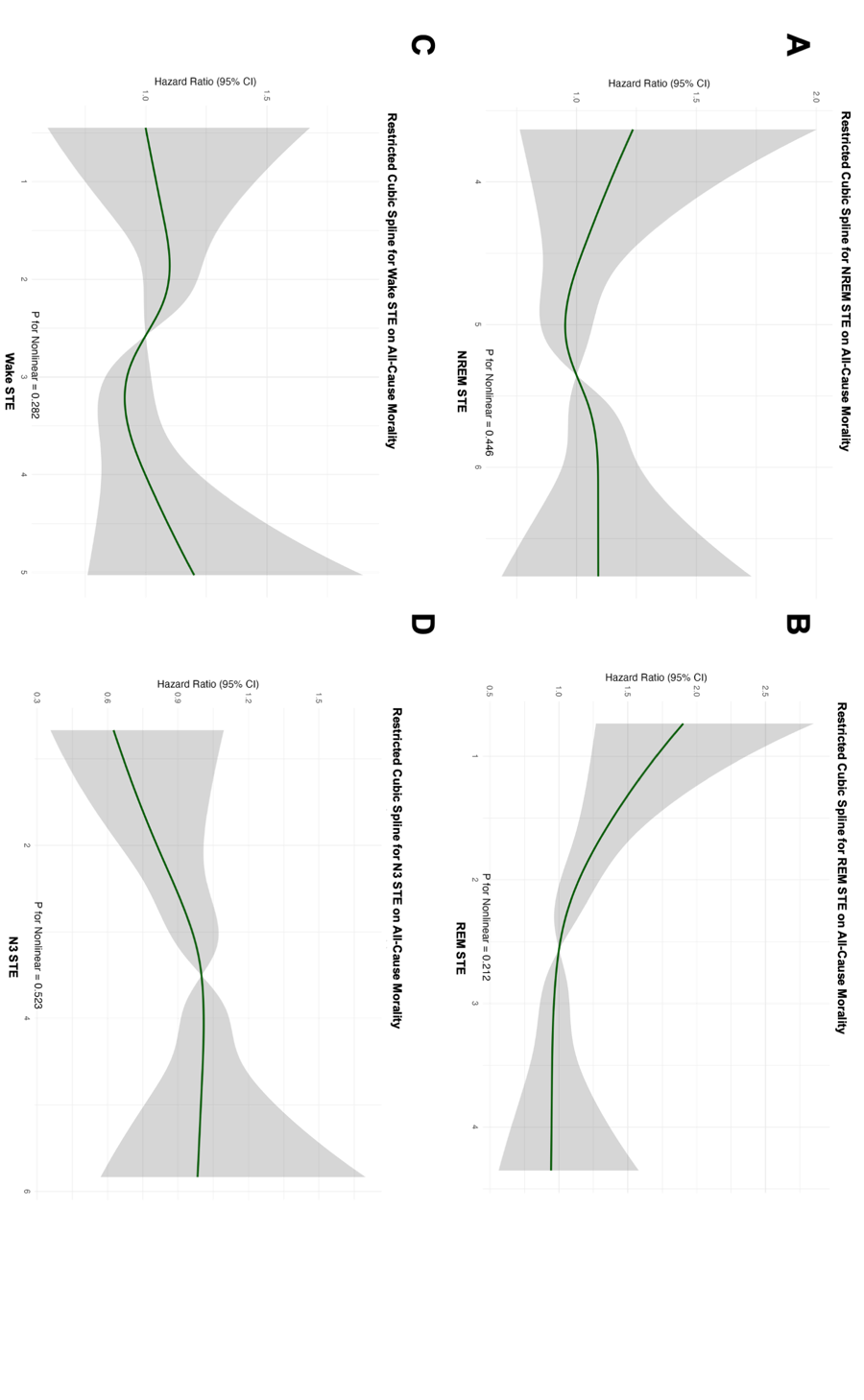

***Supplementary Figure 11: Restricted Cubic Spline (RCS) curves for fully adjusted models showing the relationship between stage-specific sleep temporal entropy (STE) and all-cause mortality:*** *Panels A–D represent: (****A****) NREM STE, (****B****) REM STE, (****C****) Wake STE, and (****D****) N3 STE. The solid green lines represent fully adjusted Hazard Ratios (HRs). Shaded areas indicate 95% confidence intervals, and p-values for nonlinearity are noted for each model.*

**2.5.2 Stage-Specific Restricted Cubic Spline Curves for CVD Mortality**

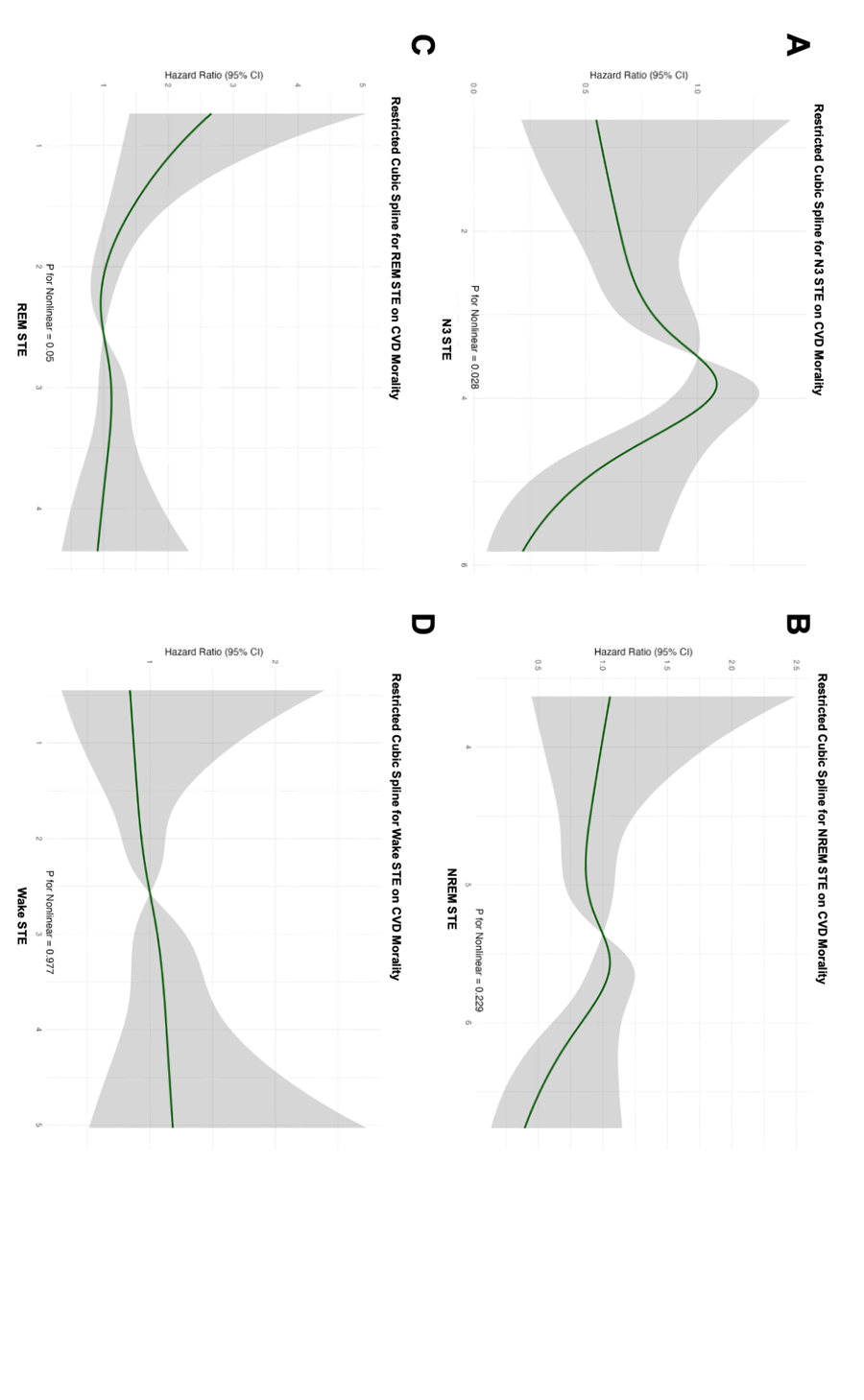

***Supplementary Figure 12: Restricted Cubic Spline (RCS) curves for fully adjusted models showing the relationship between stage-specific sleep temporal entropy (STE) and CVD mortality:*** *Panels A–D represent: (****A****) N3 STE, (****B****) NREM STE, (****C****) REM STE, and (****D****) Wake STE. The solid green lines represent fully adjusted Hazard Ratios (HRs). Shaded areas indicate 95% confidence intervals, and p-values for nonlinearity are noted for each model. Shaded areas indicate 95% confidence intervals, and p-values for nonlinearity are noted for each model.*

### 3 Dependence Plots

#### 3.1 Dependence Plots of Sleep Temporal Entropy in the SSHSC

**3.1.1 Whole-Night Dependence Plots of Sleep Temporal Entropy for Adverse Cardiometabolic Outcomes**

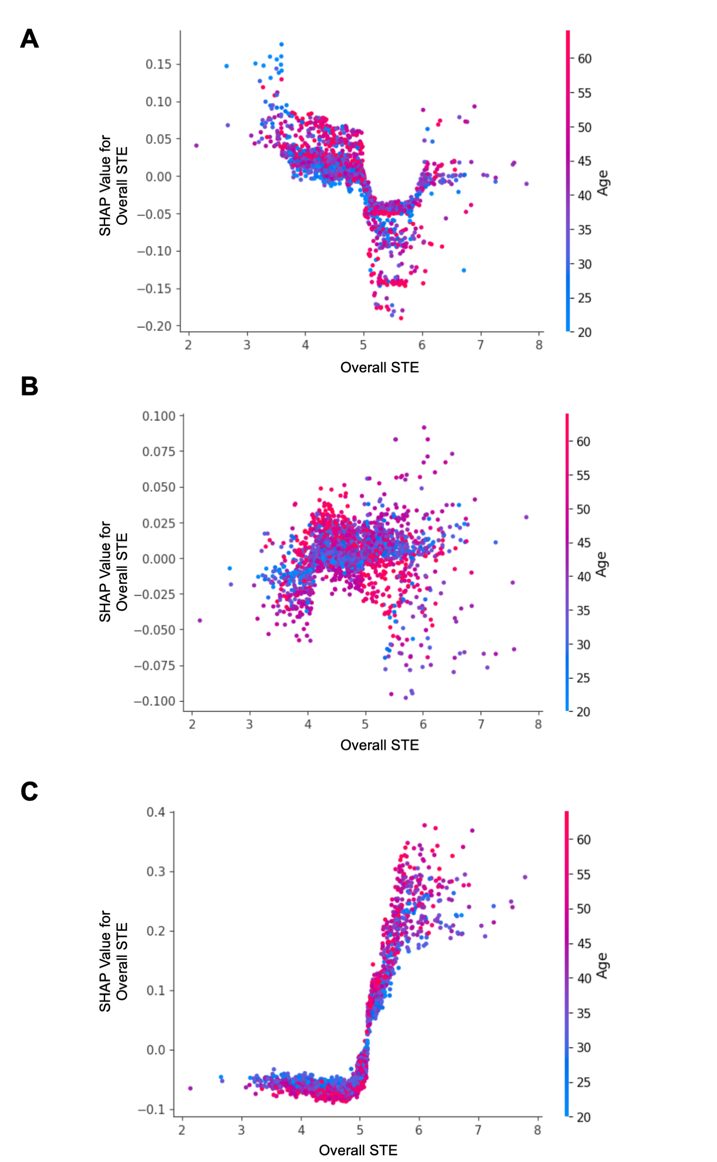

***Supplementary Figure 13: SHAP dependence plots for Overall Sleep Temporal Entropy (STE) with age represented by the color gradient, illustrating its relationship with different outcomes****: (****A****) Hypertension, (****B****) Diabetes, and (****C****) Hyperlipidemia. Each dot represents an individual participant, where the x-axis shows the OTE value, and the y-axis indicates the SHAP value, reflecting the contribution of OTE to the prediction of the outcome. Positive SHAP values indicate a higher risk of the corresponding condition, while negative values indicate a lower risk. The color gradient denotes participant age, showing the interaction effects of age and entropy on each outcome.*

**3.1.2 Stage-Specific Dependence Plots of Sleep Temporal Entropy for Hypertension**

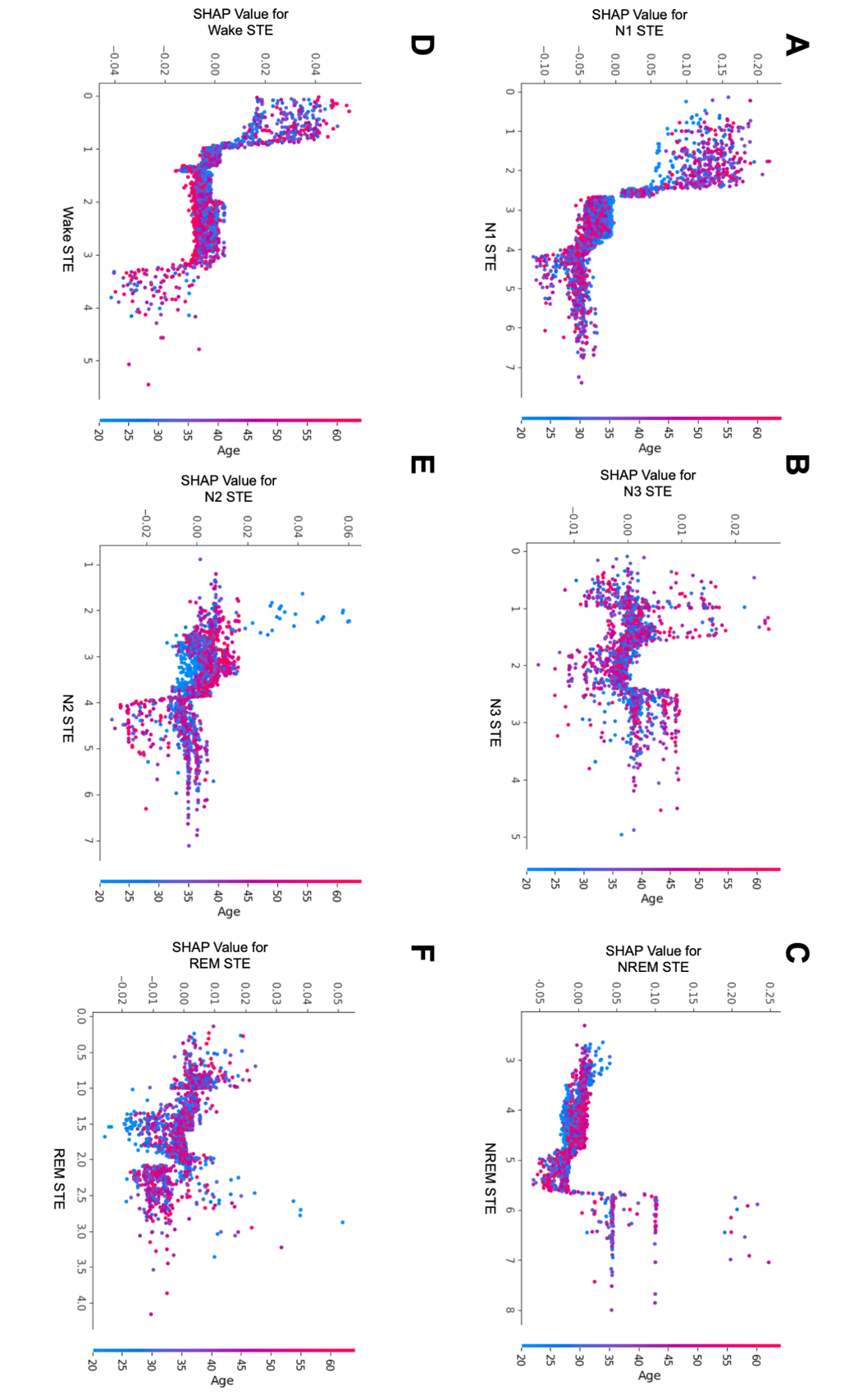

***Supplementary Figure 14: SHAP dependence plots for stage-specific temporal entropy (STE) with age represented by the color gradient, illustrating their relationship with hypertension:*** *Panels A–F correspond to: (****A****) Wake STE, (****B****) N1 STE, (****C****) N2 STE, (****D****) N3 STE, (****E****) REM STE, and (****F****) NREM STE. Each dot represents an individual participant, with the x-axis showing STE values for the corresponding sleep stage and the y-axis indicating SHAP values, reflecting the contribution of STE to the prediction of hypertension. Positive SHAP values suggest a higher risk of hypertension, while negative values indicate a lower risk. The color gradient denotes participant age, highlighting age-dependent interactions with STE in predicting hypertension.*

**3.1.3 Stage-Specific Dependence Plots of Sleep Temporal Entropy for Diabetes**

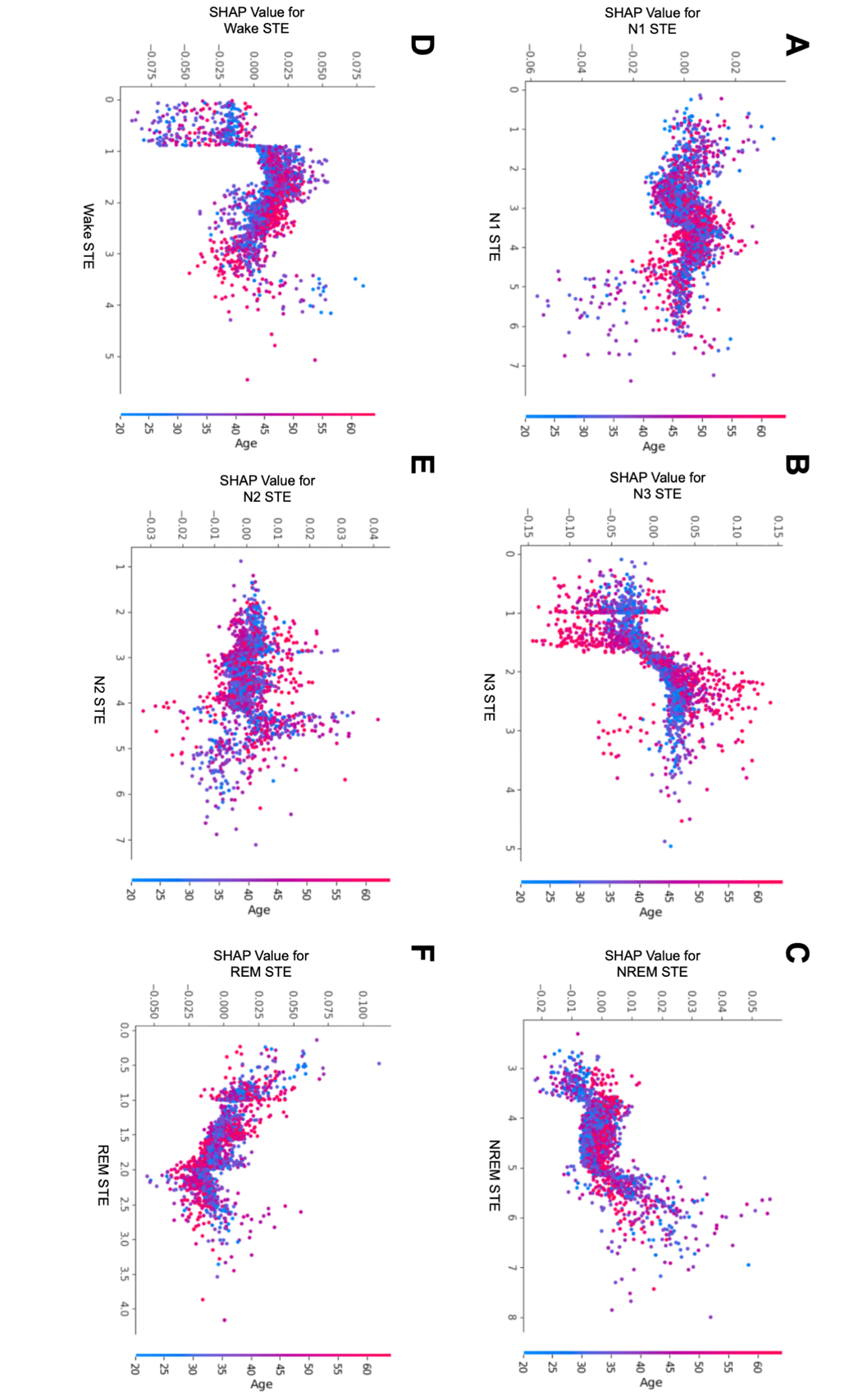

***Supplementary Figure 15: SHAP dependence plots for stage-specific temporal entropy (STE) with age represented by the color gradient, illustrating their relationship with diabetes:*** *Panels A–F correspond to: (****A****) N1 STE, (****B****) N3 STE, (****C****) NREM STE, (****D****) Wake STE, (****E****) N2 STE, and (****F****) REM STE.*

**3.1.4 Stage-Specific Dependence Plots of Sleep Temporal Entropy for Hyperlipidemia**

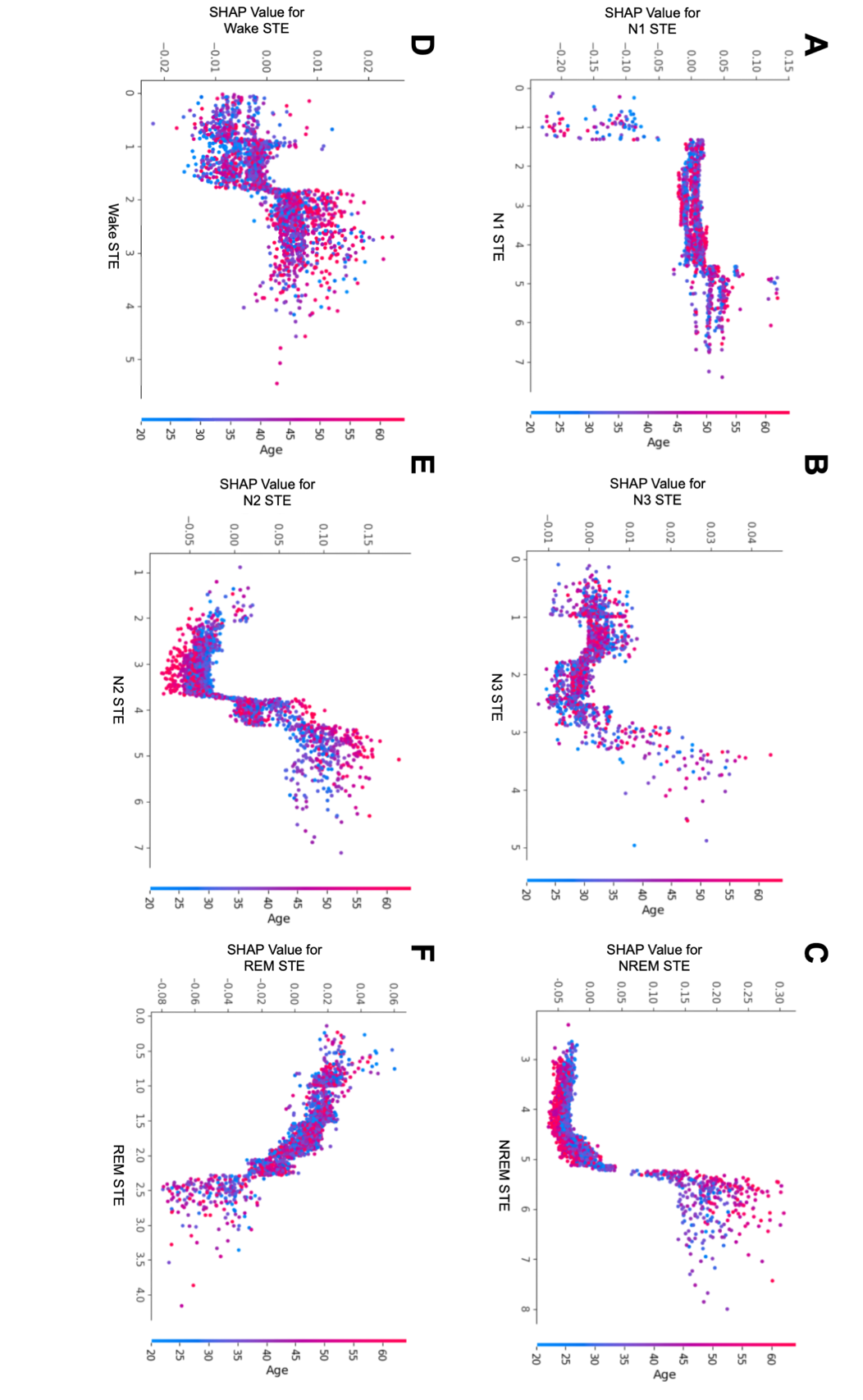

***Supplementary Figure 16: SHAP dependence plots for stage-specific temporal entropy (STE) with age represented by the color gradient, illustrating their relationship with hyperlipidemia:*** *Panels A–F correspond to: (****A****) N1 STE, (****B****) N3 STE, (****C****) NREM STE, (****D****) Wake STE, (****E****) N2 STE, and (****F****) REM STE.*

#### 3.2 Dependence Plots of Stage-Specific Temporal Entropy in the SHHS

**3.2.1 All-Cause Mortality**

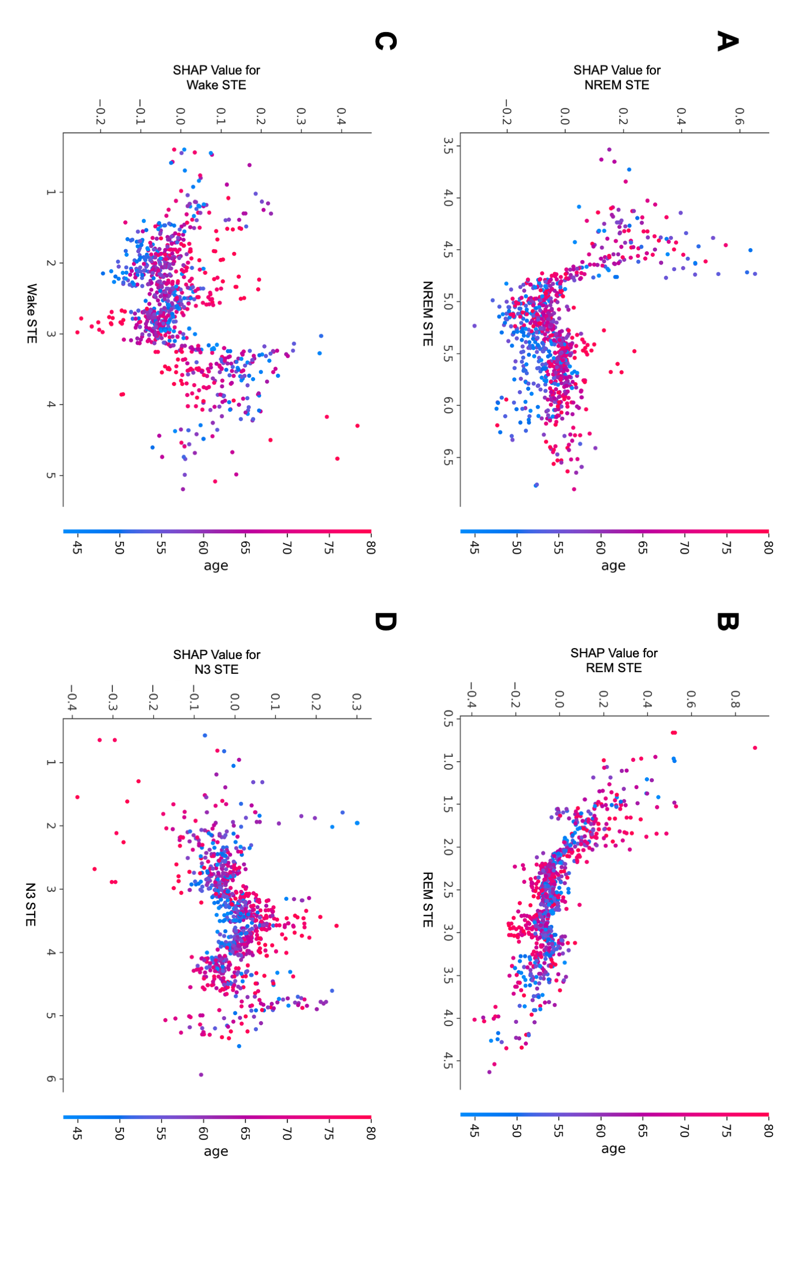

***Supplementary Figure 17: SHAP dependence plots for stage-specific sleep temporal entropy (STE) with age represented by the color gradient, illustrating their relationship with all-cause mortality in the SHHS cohort:*** *Panels A–D correspond to: (****A****) NREM STE, (****B****) REM STE, (****C****) Wake STE, and (****D****) N3 STE.*

3.2.3 **CVD Mortality**

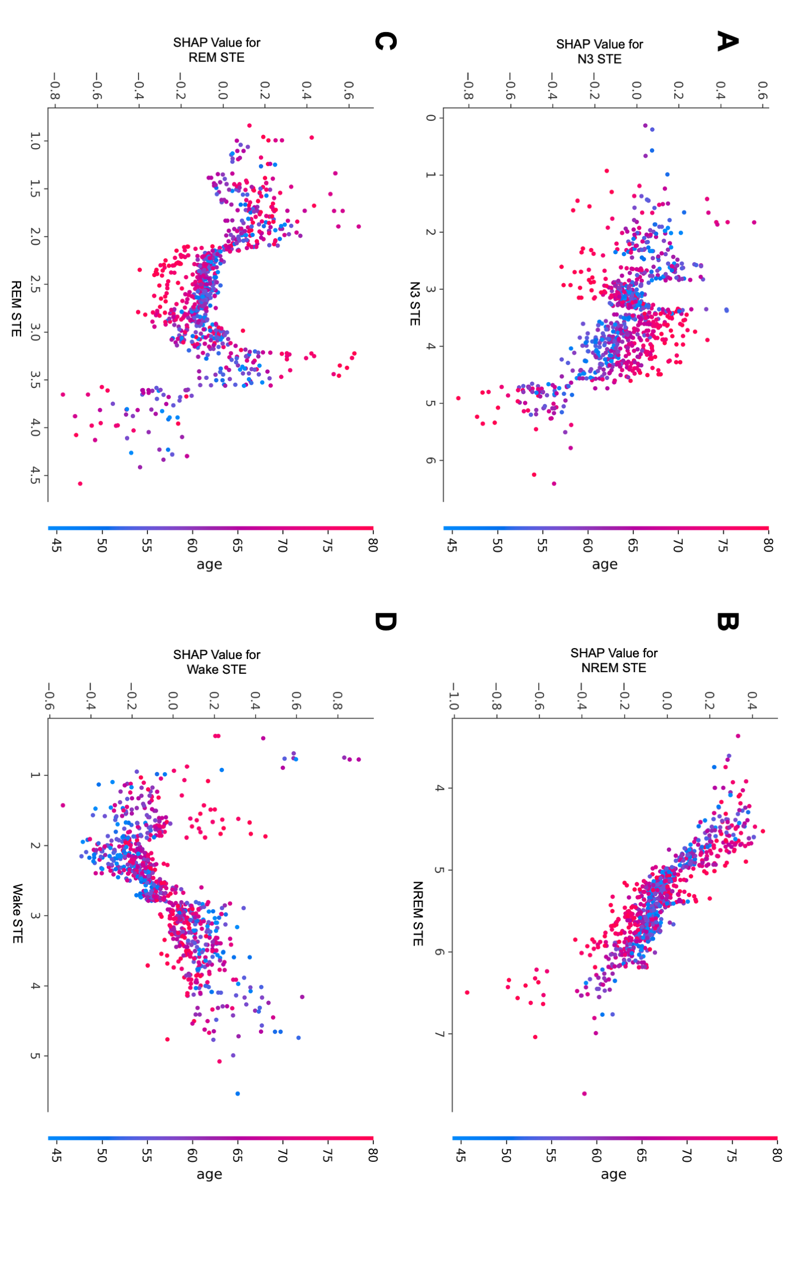

***Supplementary Figure 18: SHAP dependence plots for stage-specific sleep temporal entropy (STE) with age represented by the color gradient, illustrating their relationship with CVD mortality in the SHHS cohort:*** *Panels A–D correspond to: (****A****) N3 STE, (****B****) NREM STE, (****C****) REM STE, and (****D****) Wake STE.*
